# Continually pretrained open-weight language models can outperform traditional machine learning in EHR-based clinical prediction

**DOI:** 10.64898/2026.11.20.26368580

**Authors:** Junchao Zhou, Xinmeng Zhang, Chao Yan, Joshua Z. Ye, Mayur B. Patel, Laurie L. Novak, Abel N. Kho, Adam Wright, Bradley A. Malin, You Chen

## Abstract

Large language models (LLMs) prompted with electronic health record (EHR) data have predicted clinical outcomes less accurately than locally trained machine-learning models. We developed VEX-EHR by extending the vocabulary of a 12-billion-parameter open-weight LLM (weights publicly available) with 17,601 clinical and audit-log codes from 170,483 patients’ records at one academic medical center, continuing its pretraining on those 2019–2024 records (4.7 billion tokens) and fine-tuning it per task. AUROCs were 0.917 for next-24-hour discharge, 0.715 for 30-day readmission and 0.847 for intensive care unit acute kidney injury, against 0.868, 0.677 and 0.828 for gradient-boosted trees and 0.848, 0.650 and 0.791 for CEHR-BERT, an EHR foundation model. Continued pretraining added 0.031–0.053 AUROC on every task. With 10% of the labels, VEX-EHR exceeded both comparators using all labels for discharge, and CEHR-BERT for readmission, valuable where labels are costly. Adapted by continued pretraining, open-weight LLMs can outperform traditional machine learning in EHR-based prediction.

## Introduction

Models that predict inpatient outcomes such as discharge, readmission and acute kidney injury (AKI) are typically traditional machine-learning models, such as gradient-boosted decision trees, trained on structured electronic health record (EHR) data: diagnoses, medications, laboratory orders and, increasingly, audit-log events, the EHR’s record of which clinician opened or acted on which part of the chart and when.^1–6^ Since 2023, general-purpose large language models (LLMs), which carry broad medical knowledge, have been widely tested on the same predictions,^7–10^ yet they have not consistently outperformed these simple, locally trained models.

The most direct approach writes a patient’s record out as text and gives it to the LLM as a prompt. At our institution, GPT-4 used this way predicted next-24-hour discharge with an area under the receiver operating characteristic curve (AUROC) of 0.629, against 0.847 for gradient-boosted trees.^7^ Writing EHR events as text also makes inputs long, because each event becomes several tokens. In an earlier study at our institution, one patient-day of audit-log actions took about 38,000 characters as text, far beyond the model’s 4,096-token context window. The input therefore had to be cut to the first occurrence of each of the 10 most predictive action types. With this shortened input, a fine-tuned 70-billion-parameter LLaMA-2 predicted next-24-hour discharge with an AUROC of 0.80 when its prediction was read from the output logits, and 0.69 when read from its generated text.^11^ It remained less accurate than the gradient-boosted trees trained in the same study.^11^

EHR foundation models take another route. Models such as BEHRT, Med-BERT, CEHR-BERT and CLMBR learn a representation of a patient’s history of EHR events by self-supervision and are then fine-tuned with labeled data for each task.^12–15^ They are usually trained from scratch on one institution’s data, with a vocabulary built from its codes.^16^ They therefore learn each code only from how it appears alongside other codes, without the medical knowledge that LLMs acquire from text. Their size is also limited by the data: training from scratch makes the best use of computing at about 20 tokens per model parameter,^17^ so the 4.7 billion event tokens in this study would support a model of about 235 million parameters, one-fiftieth the size of a 12-billion-parameter LLM. Their gains over traditional models have been modest. On a public benchmark of 15 tasks, a gradient-boosted model on code counts slightly exceeded the foundation model CLMBR-T-Base when all labels were used, and pretraining helped mainly when labels were scarce.^8^

Recent work has tried to combine the two. Hegselmann et al. replaced each code with its text description, passed the text through a general LLM whose weights were left unchanged (frozen) and trained a simple classifier on its output; this matched CLMBR-T-Base, but encoding the benchmark took about 22 hours, compared with 6 minutes for CLMBR-T-Base.^8^ MedRep gives EHR foundation models such as BEHRT and Med-BERT a ready-made starting representation for each standard medical concept, built from two sources: an LLM-written description of the concept, and the concept’s links to broader and narrower concepts in the OMOP standard vocabulary (for example, a specific diagnosis and its parent category). Models that started from these representations were more accurate in external validation.^18^ EHRAdapt, the closest to our work, lets a frozen LLM read EHR events without converting them into text: a small trained add- on module (an adapter) turns each event into an input vector for the LLM, starting from an embedding of the code’s description produced by a separate biomedical language model.^19^ In all three designs, the LLM itself never learns the institution’s codes.

A different option is to make the codes part of the LLM’s own vocabulary, as is done when LLMs are taught new languages. To adapt an English-trained LLM to Chinese, for example, Chinese words are added to its vocabulary as new tokens, and the model resumes its original training task, predicting the next token, on Chinese text until it learns what the new tokens mean; this step is called continued pretraining.^20^ Because a new token starts without a learned embedding, the numerical vector through which the model reads it, the embedding must first be initialized, and how best to do so is an active area of study.^21–24^ Earlier studies added medical codes to an LLM’s vocabulary, for concepts extracted from clinical notes^25^ or for diagnosis codes,^26^ but did not evaluate the full pipeline (adding the codes, initializing them, continued pretraining and fine-tuning) for clinical prediction. Here we apply this full pipeline to EHR data.

Each clinical or audit-log code becomes one new token; its embedding is initialized from the LLM’s own representation of the code’s English description; and continued pretraining on the institution’s records teaches the model how codes follow one another in patients’ histories. Each event then costs one token, and the LLM itself is adapted: continued pretraining trains its embedding table and small low-rank adapters, about 9.5% of its parameters.

In this study, we developed VEX-EHR (vocabulary-extended large language model for EHR) by applying this approach to a 12-billion-parameter open-weight LLM with the clinical and audit-log codes of one academic medical center. We evaluated it on three inpatient tasks that span how prediction models are used: next-24-hour discharge, an operational outcome for discharge planning; 30-day readmission, an outcome after discharge and a common quality measure; and AKI onset in the intensive care unit, an acute deterioration in a small, high-risk population. Our main question was whether the adapted LLM outperforms traditional machine learning and an EHR foundation model trained on the same data, and which steps of the adaptation account for its accuracy. We also examined its accuracy in the following year, with fewer labels and across patient groups, and its cost. To our knowledge, this is the first systematic evaluation of adapting an open-weight LLM to structured EHR data through vocabulary extension, new-token initialization, continued pretraining and fine-tuning for clinical prediction.

## Results

### VEX-EHR overview and study cohort

Figure 1 summarizes the adaptation pipeline and the analyses. VEX-EHR adds the institution’s clinical and audit-log codes to the vocabulary of a general-purpose LLM (Gemma-4 12B), initializes each new code token from the code’s text description and continues pretraining on the institution’s event sequences. It is then fine-tuned for each task: the adapted LLM encodes the patient’s events in the 24 hours before the prediction time, one token per event, and a task-specific prediction head estimates the probability of the outcome.

**Figure 1.**
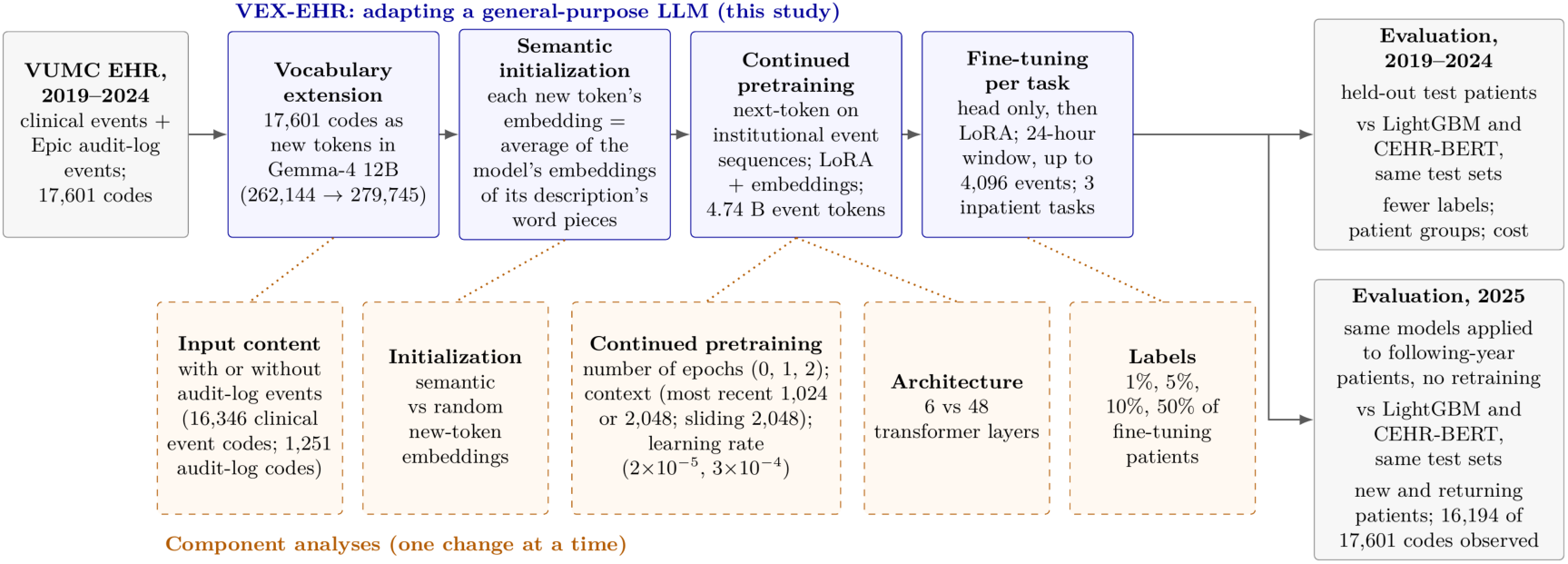
Study design. Clinical and Epic audit-log events from one health system (2019–2024; 17,601 vocabulary tokens: 17,597 codes and 4 special tokens; Supplementary Table S1) are added to a general-purpose LLM’s vocabulary as new tokens (vocabulary extension), and each new token is initialized from the model’s own representation of the code’s description (semantic initialization). The model is then continued-pretrained on the institution’s event sequences, training low-rank adapters and the embedding table, and fine-tuned for each task. Component analyses (dashed boxes) change one step at a time: the input content (with or without audit-log events); the initialization of the new tokens (from descriptions or at random); continued *pretraining (none, one or two epochs; a fixed window of the most recent 1,024 or 2,048 events of each record, or 2,048-token windows sliding over the record, with a fine-tuning input of the most recent 1,024, 2,048 or 4,096 tokens, respectively; a learning rate of 2×10^−5^ or 3×10^−4^) and transformer depth (6 or 48 layers); and the share of fine-tuning labels. The adapted model (VEX-EHR) is evaluated on held-out 2019–2024 test patients, against LightGBM and CEHR-BERT trained on the same data, and, separately, on a 2025 cohort without retraining, in which 16,194 of the 17,601 codes occur. LLM, large language model; LoRA, low-rank adaptation*.

The cohort comprised 170,483 patients for pretraining, 5,383 for fine-tuning and 3,590 for testing, with no patient appearing in more than one set (Table 1). Each record is a time-ordered sequence of events, each represented by one of 17,601 distinct codes that form the vocabulary: clinical events (diagnoses, medications, procedures, laboratory orders, demographics and encounters) and Epic audit-log actions, which record which part of the chart a clinician opened or acted on. Audit-log codes constituted 7% of the vocabulary but accounted for 89.9% of event occurrences in the full-history pretraining corpus.

**Table 1.** Prediction tasks, cohorts and splits. The test sets are identical, instance for instance, for all models. Fine-tuning training/validation counts are for seed 42; the split moves with the fine-tuning seed and the test set does not. 2025: the following year, scored without retraining.

|  | Discharge (next 24 h) | 30-day readmission | ICU AKI onset (48 h) |
|---|---|---|---|
| Prediction time | 14:00 daily, from day 2 | at discharge | every 6 h in ICU |
| Input window | 24 h before prediction | 24 h before prediction | 24 h before prediction |
| *Training data, 2019–2024* |  |  |  |
| Continued pretraining (all tasks) | 170,483 patients; 4.7 billion event tokens |  |  |
| Fine-tuning pool, instances | 38,290 | 8,360 | 8,059 |
| training / validation | 34,447 / 3,843 | 7,525 / 835 | 7,256 / 803 |
| *Test set, 2019–2024* |  |  |  |
| Test instances | 26,302 | 5,471 | 5,226 |
| Test patients | 3,345 | 3,537 | 392 |
| Test instances per patient, mean | 7.9 | 1.5 | 13.3 |
| Test instances with the outcome | 12.5% (3,292) | 10.9% (594) | 10.9% (571) |
| Input tokens retained at 4,096 | 97.7% | 99.8% | 92.8% |
| Age, median (IQR) | 53 (33–67) | 53 (34–68) | 60 (45–69) |
| Female, % | 57.0 | 56.2 | 41.3 |
| Race, %: White | 76.9 | 77.1 | 77.6 |
| non-White | 23.1 | 22.9 | 22.4 |
| *Test set, 2025 (the following year; no retraining)* |  |  |  |
| Test instances | 256,652 | 55,824 | 35,601 |
| Test patients | 40,499 | 42,234 | 3,264 |
| Test instances per patient, mean | 6.3 | 1.3 | 10.9 |
| Test instances with the outcome | 10.6% (27,263) | 12.1% (6,756) | 17.0% (6,035) |
| Input tokens retained at 4,096 | 96.1% | 99.6% | 86.1% |
| Age, median (IQR) | 55 (35–70) | 55 (35–70) | 60 (47–69) |
| Female, % | 58.5 | 58.1 | 41.5 |
| Race, %: White | 76.0 | 76.2 | 77.6 |
| non-White | 24.0 | 23.8 | 22.4 |

Pretraining: 170,483 patients, 2019–2024; a patient admitted in more than one calendar year contributes one record per year, giving 209,764 records; fine-tuning pool 5,383 patients; test 3,590 patients; the three sets are disjoint by patient. Discharge instances whose input window contains a discharge-day billing code (CPT 99238/99239) are excluded (69,864 → 64,592 labeled instances before the patient-level split). AKI is defined by KDIGO creatinine criteria; instances after the first onset in a stay, and stays ending before the 48-hour outcome window without AKI, are excluded. “Input tokens retained” is the share of input-window events inside the 4,096-token context (2019–2024: rows of fine-tuning and test patients; 2025: all rows).
Patient characteristics are for the patients with a test instance: age at the first prediction (calendar year minus year of birth); sex and race from the OMOP person record; non-White includes every other recorded race and unknown or declined race, as in the race analysis. ICU, intensive care unit; AKI, acute kidney injury.

We evaluated three prediction tasks, each predicting a binary outcome at a fixed time (one prediction instance): discharge within the next 24 hours, predicted daily at 14:00 from the second hospital day; readmission within 30 days, predicted at discharge; and acute kidney injury (AKI) onset within 48 hours, predicted every 6 hours in the intensive care unit (ICU). Across the 2019–2024 test sets, 10.9–12.5% of the instances had the outcome. Because discharge and AKI are predicted repeatedly during a stay and readmission once per stay, one patient can contribute several instances: on average 7.9 per test patient for discharge, 1.5 for readmission and 13.3 for AKI; paired comparisons therefore resample patients rather than instances (Methods).

### Accuracy compared with established models

Table 2 and Figure 2a, c and e compare VEX-EHR with two established models trained on the same data (Methods): (1) LightGBM, a gradient-boosted decision-tree model based on counts of clinical and audit-log codes,^2^ and (2) CEHR-BERT, a transformer EHR foundation model that encodes the timing of events and is pretrained from scratch on this study’s pretraining patients and fine-tuned for each task.^14^ For readmission and AKI, VEX-EHR’s values are means over three fine-tuning seeds; discharge was fine-tuned once (Methods). For next-24-hour discharge, VEX-EHR achieved an AUROC of 0.917 (95% CI 0.912–0.923), compared with 0.868 (0.861–0.874) for LightGBM and 0.848 (0.841–0.855) for CEHR-BERT. For 30-day readmission, its AUROC was 0.715 (0.694–0.734), compared with 0.677 (0.653–0.699) and 0.650 (0.627–0.672), and for ICU AKI onset 0.847 (0.831–0.864), compared with 0.828 (0.810–0.847) and 0.791 (0.772–0.811). VEX-EHR also had a higher area under the precision-recall curve (AUPRC) than both comparators on every task: 0.711, 0.266 and 0.473 for discharge, readmission and AKI, against 0.582, 0.260 and 0.444 for LightGBM and 0.605, 0.217 and 0.357 for CEHR-BERT (Table 2).

**Figure 2.**
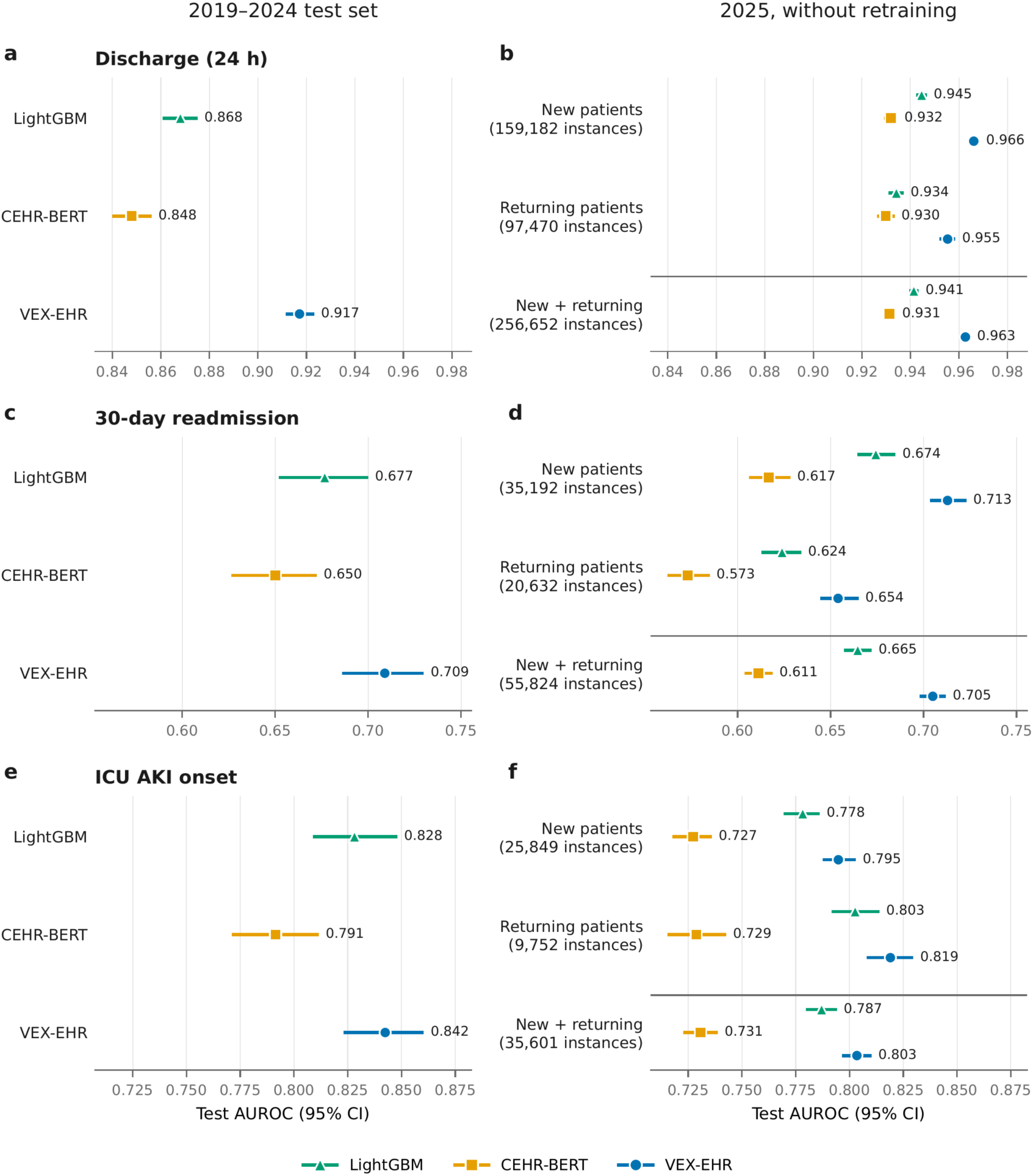
Discrimination of VEX-EHR and established models. Test AUROC with 95% bootstrap intervals for LightGBM, CEHR-BERT and VEX-EHR (fine-tuning seed 42), one row per task (a, b, discharge; c, d, readmission; e, f, AKI); the two panels of a row share the horizontal axis. a, c, e, the 2019–2024 test sets. b, d, f, the 2025 cohort scored without retraining, all models on the same instances: new patients (absent from all 2019–2024 data), returning patients, and both *together (new + returning, below the line). AUROC values are printed next to the points; the 2019–2024 and new + returning values, and the means over three fine-tuning seeds for readmission and AKI, are in Table 2. Intervals narrower than the marker are hidden by it. AKI, acute kidney injury; AUROC, area under the receiver operating characteristic curve*.

**Table 2.** Test performance on the 2019–2024 held-out test sets and, without retraining, in 2025. Values carry 95% percentile bootstrap intervals over test instances (1,000 resamples, seed 42). The highest value in each column of each block is in bold.

| Model | AUROC (95% CI) | AUPRC (95% CI) |
|---|---|---|
| ***2019–2024 test set*** |  |  |
| **Next-24-hour discharge** (*n* = 26,302; 3,345 patients) |  |  |
| LightGBM<sup>a</sup> | 0.8680 (0.8613–0.8745) | 0.5819 (0.5642–0.5983) |
| CEHR-BERT<sup>a</sup> | 0.8480 (0.8405–0.8555) | 0.6055 (0.5889–0.6240) |
| VEX-EHR-Lite (Gemma-4 12B), seed 42 | 0.9006 (0.8948–0.9064) | 0.6853 (0.6672–0.7021) |
| VEX-EHR (Gemma-4 12B), seed 42 | **0.9172 (0.9121–0.9225)** | **0.7111 (0.6947–0.7274)** |
| **30-day readmission** (*n* = 5,471; 3,537 patients) |  |  |
| LightGBM<sup>a</sup> | 0.6767 (0.6527–0.6992) | 0.2601 (0.2283–0.2963) |
| CEHR-BERT<sup>a</sup> | 0.6501 (0.6273–0.6716) | 0.2172 (0.1907–0.2479) |
| VEX-EHR-Lite (Gemma-4 12B), seed 42 | 0.7090 (0.6864–0.7298) | **0.2706 (0.2384–0.3053)** |
| VEX-EHR (Gemma-4 12B), seed 42 | 0.7090 (0.6870–0.7291) | 0.2587 (0.2284–0.2939) |
| VEX-EHR (Gemma-4 12B), mean of 3 seeds | **0.7153 (0.6937–0.7340)** | 0.2664 (0.2365–0.3027) |
| **ICU AKI onset** (*n* = 5,226; 392 patients) |  |  |
| LightGBM<sup>a</sup> | 0.8282 (0.8096–0.8473) | 0.4445 (0.3994–0.4880) |
| CEHR-BERT<sup>a</sup> | 0.7914 (0.7719–0.8108) | 0.3567 (0.3187–0.3971) |
| VEX-EHR-Lite (Gemma-4 12B), seed 42 | 0.8078 (0.7872–0.8271) | 0.3715 (0.3338–0.4086) |
| VEX-EHR (Gemma-4 12B), seed 42 | 0.8424 (0.8237–0.8596) | 0.4716 (0.4299–0.5123) |
| VEX-EHR (Gemma-4 12B), mean of 3 seeds | **0.8475 (0.8306–0.8640)** | **0.4732 (0.4328–0.5121)** |
| ***2025, without retraining*** |  |  |
| **Next-24-hour discharge** (*n* = 256,652; 40,499 patients) |  |  |
| LightGBM | 0.9414 (0.9401–0.9428) | 0.7150 (0.7097–0.7203) |
| CEHR-BERT | 0.9314 (0.9296–0.9331) | 0.7666 (0.7617–0.7714) |
| VEX-EHR-Lite (Gemma-4 12B), seed 42 | **0.9738 (0.9729–0.9747)** | **0.8535 (0.8492–0.8575)** |
| VEX-EHR (Gemma-4 12B), seed 42 | 0.9627 (0.9612–0.9640) | 0.8382 (0.8336–0.8427) |
| **30-day readmission** (*n* = 55,824; 42,234 patients) |  |  |
| LightGBM | 0.6646 (0.6582–0.6712) | 0.2237 (0.2148–0.2325) |
| CEHR-BERT | 0.6112 (0.6046–0.6179) | 0.1750 (0.1686–0.1814) |
| VEX-EHR-Lite (Gemma-4 12B), seed 42 | 0.6866 (0.6802–0.6931) | 0.2413 (0.2318–0.2510) |
| VEX-EHR (Gemma-4 12B), seed 42 | **0.7050 (0.6988–0.7112)** | **0.2572 (0.2477–0.2673)** |
| VEX-EHR (Gemma-4 12B), mean of 3 seeds | 0.7032 (0.6971–0.7092) | 0.2551 (0.2458–0.2649) |
| **ICU AKI onset** (*n* = 35,601; 3,264 patients) |  |  |
| LightGBM | 0.7870 (0.7806–0.7934) | 0.4417 (0.4293–0.4544) |
| CEHR-BERT | 0.7308 (0.7234–0.7380) | 0.3583 (0.3457–0.3708) |
| VEX-EHR-Lite (Gemma-4 12B), seed 42 | 0.7604 (0.7538–0.7671) | 0.3863 (0.3733–0.3992) |
| VEX-EHR (Gemma-4 12B), seed 42 | 0.8034 (0.7973–0.8095) | 0.4549 (0.4412–0.4689) |
| VEX-EHR (Gemma-4 12B), mean of 3 seeds | **0.8046 (0.7986–0.8103)** | **0.4593 (0.4465–0.4728)** |

<sup>a</sup>Trained on the same fine-tuning sets and evaluated on the same test instances (Methods; Supplementary Section S4). Differences from VEX-EHR were tested in pairs (patient-clustered bootstrap; Results). Mean of 3 seeds: AUROC or AUPRC averaged over fine-tuning seeds 42, 142 and 242 with continued pretraining fixed, its interval from the same resamples (test-set sampling only); discharge was fine-tuned at seed 42 only; per-seed values are in Supplementary Table S14. 2025: the same models applied to every 2025 instance, all models on the same instances. Discharge: VEX-EHR and CEHR-BERT scored the same 26,300 of the 26,302 test instances (two negative instances without events in the input window); VEX-EHR-Lite scored all 26,302. VEX-EHR-Lite: the lower-cost configuration (continued pretraining on the most recent 2,048 events of each record, two epochs at the lower learning rate; fine-tuning on 2,048 tokens; Methods). AKI, acute kidney injury; AUPRC, area under the precision-recall curve; AUROC, area under the receiver operating characteristic curve.

In paired comparisons on identical test instances (patient-clustered bootstrap; Supplementary Table S4), VEX-EHR’s AUROC exceeded that of LightGBM by 0.049 (95% CI 0.043–0.056) for discharge and by 0.039 on average over the three fine-tuning seeds for readmission, with the 95% CI excluding zero at every seed. For AKI, the average difference from LightGBM, 0.019, was not statistically significant (the 95% CI included zero at every seed; p = 0.07–0.40). VEX-EHR’s AUROC exceeded that of CEHR-BERT by 0.069 (95% CI 0.062–0.077) for discharge, and on average by 0.065 for readmission and 0.056 for AKI, with the 95% CI excluding zero at every seed.

### Contribution of each adaptation component

Figure 3 quantifies each component of the adaptation (numbers in Supplementary Table S6). Because continued pretraining of VEX-EHR is costly, the component analysis started from a lower-cost reference configuration: two epochs of continued pretraining over the most recent 1,024 events of each record, and fine-tuning on a 1,024-token input. This reference reached AUROCs of 0.882 for discharge, 0.689 for readmission and 0.814 for AKI. Each contrast changed one component, repeated continued pretraining and fine-tuning, and compared test AUROC with the reference on the same test instances. Unless stated otherwise, the differences below have 95% CIs excluding zero. Continued pretraining was the only component that improved all three tasks: adding it increased AUROC by 0.051 for discharge, 0.031 for readmission and 0.053 for AKI.

**Figure 3.**
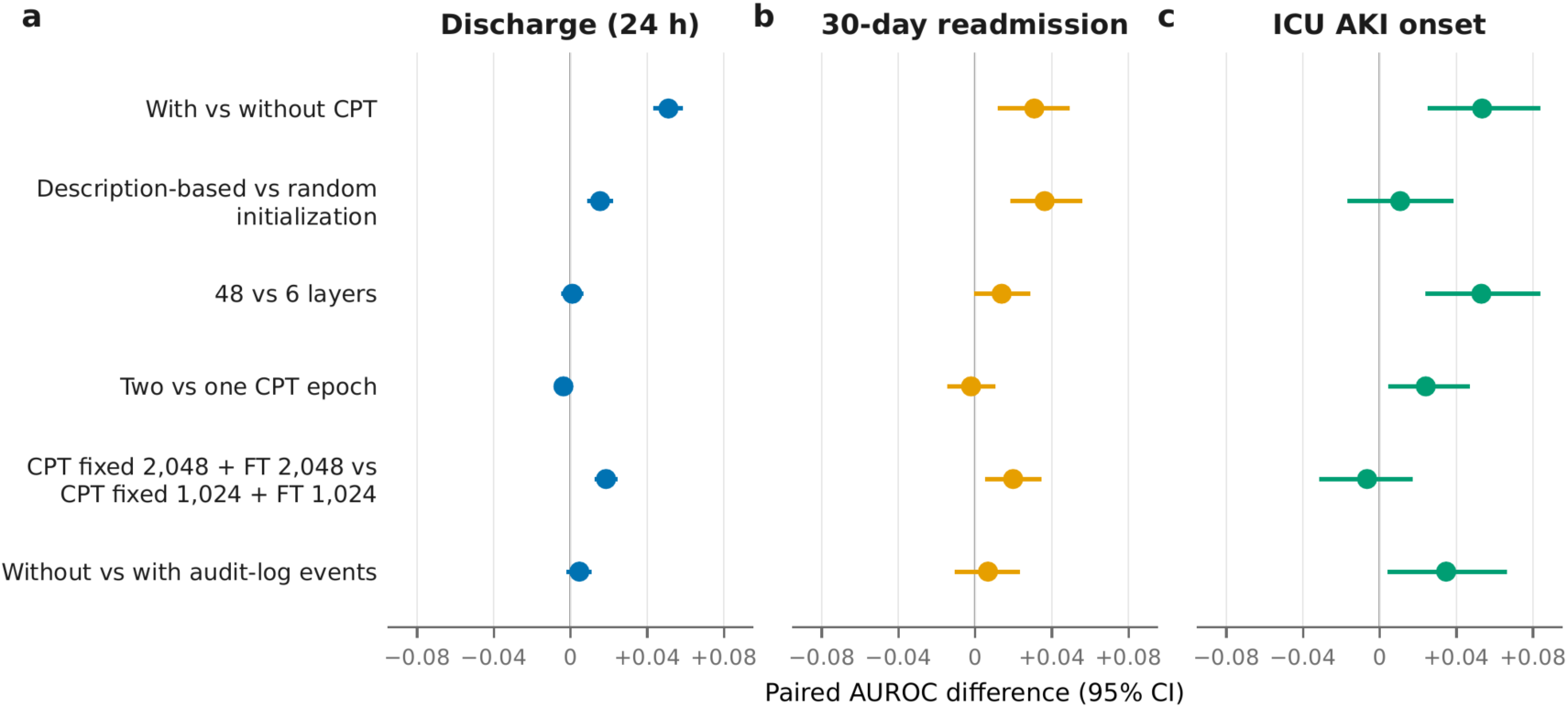
Contribution of each adaptation component. One column per task: a, discharge; b, readmission; c, AKI. Each row “A vs B” gives the test AUROC of A minus that of B, paired on the same test instances, with a patient-clustered 95% bootstrap interval (2,000 draws); an interval that does not cross the zero line excludes zero. One component of the reference configuration (CPT fixed 1,024, two epochs at 2×10^−5^; FT 1,024) is changed at a time. CPT, continued pretraining; FT, fine-tuning; CPT fixed n uses the most recent n events of each record and FT n uses the most recent n tokens of the 24-hour input. Fine-tuning seed 42. Numbers are in Supplementary Table S6. Intervals narrower than the marker are hidden by it.

Initializing the new tokens from their descriptions, rather than at random, added 0.015 on discharge and 0.036 on readmission, but not significantly on AKI (0.011, 95% CI −0.016 to 0.037). It also lowered the pretraining loss, which measures how poorly the model predicts the next event in a record, from 1.953 to 1.886.

Using all 48 transformer layers of the backbone instead of only the first 6 increased AUROC on readmission (by 0.014) and AKI (by 0.053) but not on discharge (+0.001, 95% CI −0.004 to 0.006), although the 6-layer model had the worst pretraining loss of any configuration (2.340). Pretraining loss therefore did not indicate which configuration would predict clinical outcomes best (Supplementary Section S5).

How long the model was trained, and how much of each record it read, affected the tasks differently. A second epoch of continued pretraining improved only AKI (by 0.024), whereas a longer input (2,048 instead of 1,024 tokens, in both continued pretraining and fine-tuning) improved discharge (by 0.018) and readmission (by 0.020) but not AKI; we call this 2,048-token configuration VEX-EHR-Lite. Continued pretraining over each record’s full history, with a 4,096-token input and a higher learning rate for a shorter run, gave the main model, VEX-EHR. At the same fine-tuning seed, VEX-EHR was more accurate than VEX-EHR-Lite on discharge (by 0.017) and AKI (by 0.035) and as accurate on readmission (Supplementary Section S6).

Removing audit-log events from both pretraining and task inputs did not significantly change discharge (+0.005) or readmission (+0.007) but improved AKI by 0.035. This comparison measures a trade-off within a fixed input length, not the information that audit-log events carry. Because this configuration reads at most the 1,024 most recent events, removing audit-log events let more clinical history into the input: audit-log events made up 84% of the events in AKI inputs, and removing them raised the median number of clinical events per prediction from 136 to 282 and lowered the share of predictions without a creatinine-related laboratory order from 56% to 22% (Supplementary Table S8). In an occlusion analysis of VEX-EHR-Lite, clinical events made up 17% of AKI inputs but accounted for 69% of the change in predictions when events of one type were deleted, and audit-log events for the remaining 31% (Supplementary Section S8).

### Temporal validation in 2025

Applied without retraining to the 2025 cohort, VEX-EHR reached AUROCs of 0.963 for discharge (256,652 instances), 0.703 for readmission (55,824 instances) and 0.805 for AKI (35,601 instances); readmission and AKI values are means over three fine-tuning seeds, and discharge was fine-tuned once (Figure 2b, d and f; Supplementary Section S9). Compared with the 2019–2024 test sets, accuracy was higher for discharge (0.917 in 2019–2024), similar for readmission (0.715) and lower for AKI (0.847); on AKI, LightGBM (0.828 to 0.787) and CEHR-BERT (0.791 to 0.731) declined by similar or larger amounts (Table 2). Accuracy in new patients, absent from all 2019–2024 data, was higher than in returning patients for discharge (0.966 against 0.955; difference 0.011, 95% CI 0.008–0.014) and readmission (0.711 against 0.654; 0.057, 0.042–0.072); for AKI it was lower (0.795 against 0.821), but the difference was not statistically significant (−0.026, −0.058 to 0.007).

On the same 2025 instances, VEX-EHR was more accurate than CEHR-BERT on every task (by 0.031, 0.092 and 0.074 for discharge, readmission and AKI) and than LightGBM on discharge (by 0.021) and readmission (by 0.039), with 95% CIs excluding zero in all, new and returning patients and at every fine-tuning seed. On AKI, its advantage over LightGBM (0.018 on average) was not significant at every seed: the 95% CI included zero at one of the three seeds (Supplementary Table S11). Compared with VEX-EHR-Lite, VEX-EHR was more accurate in 2025 on AKI at every seed (by 0.039 on average) and less accurate on discharge (by 0.011; one seed), with 95% CIs excluding zero; on readmission it was more accurate at two of the three seeds (by 0.018 on average) (Supplementary Table S7).

### Learning from fewer labels

We fine-tuned the continued-pretrained model on 1%, 5%, 10% and 50% of the fine-tuning patients, at three fine-tuning seeds (50%: seed 42 only; Figure 4; Supplementary Section S10). VEX-EHR’s AUROC was above that of LightGBM and CEHR-BERT trained on the same patients at every fraction on every task (AKI at 1%, with 4 patients, could not be estimated). In paired comparisons with LightGBM trained on the same patients, the difference was significant at every fraction of discharge, at 10% and 50% of readmission and at 5% of AKI, but not at 5% of readmission or at 10% and 50% of AKI. With 5% of the discharge labels (about 1,900 labeled instances), VEX-EHR reached an average AUROC of 0.862, above CEHR-BERT trained on all labels (0.848), and with 10% (about 4,300 instances) 0.888, above LightGBM trained on all labels (0.868). On readmission, 10% of the labels (about 880 instances) gave 0.659, above CEHR-BERT with all labels (0.650), and 50% (about 4,200 instances) gave 0.702, above LightGBM with all labels (0.677). On AKI, 50% of the labels (about 4,300 instances) gave 0.828, above CEHR-BERT with all labels (0.791) and equal to LightGBM with all labels (0.828).

**Figure 4.**
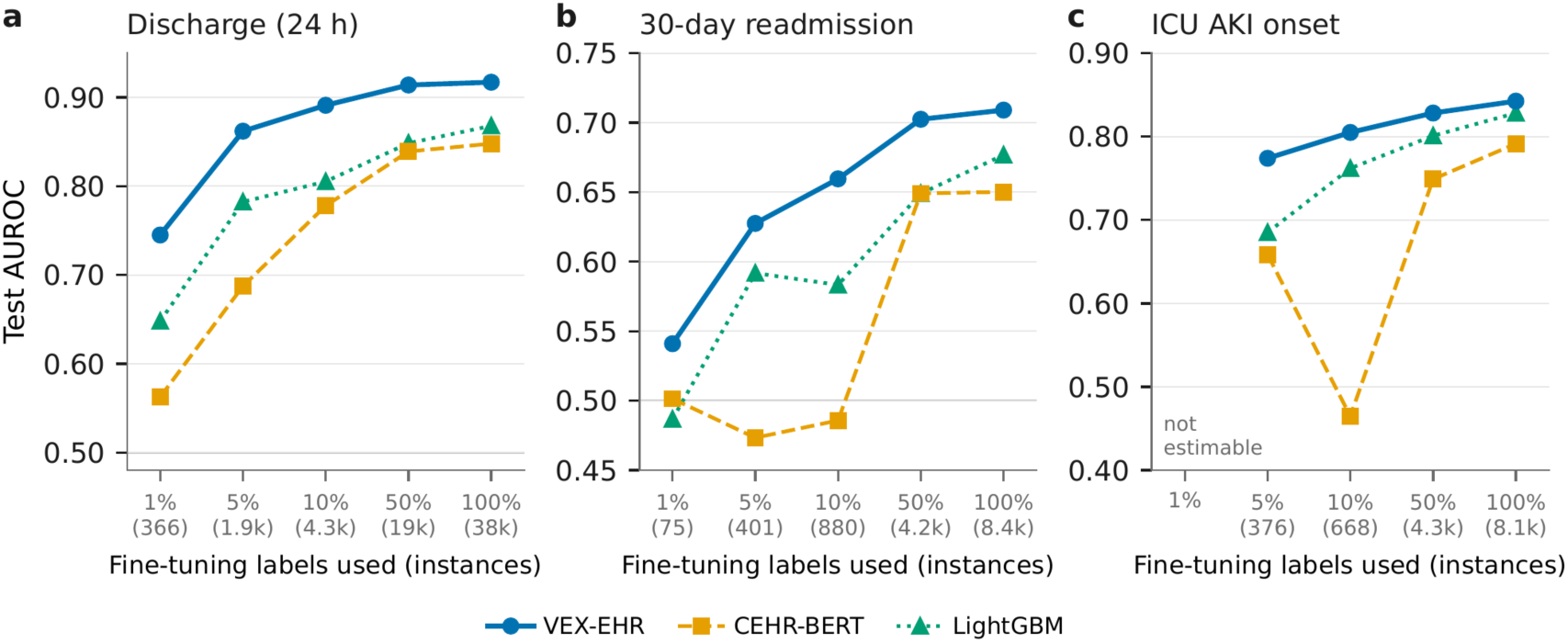
Test AUROC against the fraction of fine-tuning labels used, one panel per task (a, discharge; b, readmission; c, AKI); in parentheses, the number of labeled fine-tuning instances (training and validation). All three models were fine-tuned on the same patient subsets (fine-tuning seed 42; the means over three seeds quoted in the text are in Supplementary Table S12); 100% is the full fine-tuning set (Table 2). At some small fractions (discharge 1%; readmission 1%, 5% and 10%; AKI 10%) CEHR-BERT’s predictions were nearly constant (standard deviation of the predicted probabilities below 0.005), giving AUROCs near 0.5; repeated at fine-tuning seeds 142 and 242, all 15 runs at these fractions gave AUROCs of 0.46–0.57, 13 of them with nearly constant predictions. AKI at 1% (4 patients) is not estimable. VEX-EHR and CEHR-BERT (all three seeds) and LightGBM values are in Supplementary Table S12, and AUPRC in Supplementary Section S10.

### Stability and calibration

For readmission and AKI, the three fine-tuning seeds gave test AUROCs within 0.02 of each other (Supplementary Section S11). Before recalibration, the model was poorly calibrated: its raw risk estimates were higher than the observed rates, because positive cases, being less common, were given extra weight during training. Refitting the estimates on the validation data (logistic recalibration) brought them close to the observed rates: the average gap between predicted and observed risk (the expected calibration error) fell to 0.010–0.033. This step changes the risk values but not the order of the patients, so AUROC is unchanged (Supplementary Section S12). Although AUROC varied little between fine-tuning seeds, the decision threshold did not. The threshold chosen on each run’s validation set (maximizing sensitivity plus specificity, Youden’s index) differed between seeds; applied to the 2025 AKI cohort, the three seeds’ thresholds would have flagged 18%, 44% and 39% of instances as high risk. A threshold for clinical use would therefore need to be set on local data (Supplementary Section S13).

### Performance across patient groups

Patients recorded as a race other than White, or whose race was unknown or declined, were grouped as non-White, following a previous study;^7^ they accounted for 23% of the 2019–2024 test patients and 24% of the 2025 cohort. VEX-EHR’s AUROC was not significantly lower in non-White than in White patients for any task, in the 2019–2024 test set or in 2025 (all, new and returning patients), at any fine-tuning seed; where the difference was significant, AUROC was higher in non-White patients (discharge in 2025, 0.971 against 0.960; readmission in 2025, 0.740 against 0.691, at every seed; Supplementary Section S13). By sex and age (2019–2024 test set, seed 42; exploratory), discharge AUROC was at least 0.895 in every group, whereas readmission AUROC was lower in men (0.669) and in patients aged 65 or older (0.650) than in women (0.738) and in patients younger than 45 (0.763), and AKI AUROC was lowest in patients aged 65 or older (0.793, against 0.886 in patients younger than 45).

### Training cost

Adaptation took 940 GPU-hours on NVIDIA H100 GPUs (Supplementary Table S2): 730 for continued pretraining, a one-time step shared by all tasks, and 210 for fine-tuning one model per task (153 for discharge, 23 for readmission and 34 for AKI). Continued pretraining updated 1.14 billion parameters (the low-rank adapters and the embedding table) and fine-tuning 66.8 million (0.55% of the model). The lower-cost VEX-EHR-Lite configuration required 250 GPU-hours, 114 for continued pretraining and 136 for fine-tuning, with a small loss of accuracy: at the same fine-tuning seed, its AUROC was 1.8% lower than that of VEX-EHR on discharge and 4.1% lower on AKI, and equal on readmission (Supplementary Table S6). A 6-layer model trained 3.7 times as fast per step as the full 48-layer model.

### How the new tokens changed

Figure 5 maps the vocabulary in two dimensions; distances and neighbors were measured in the original 3,840-dimensional embedding space (Supplementary Section S14). After semantic initialization, codes lay mostly near codes of their own type (diagnosis, medication, procedure, laboratory or audit-log; 97.4% of each code’s 10 nearest codes were of the same type; Figure 5a).

**Figure 5.**
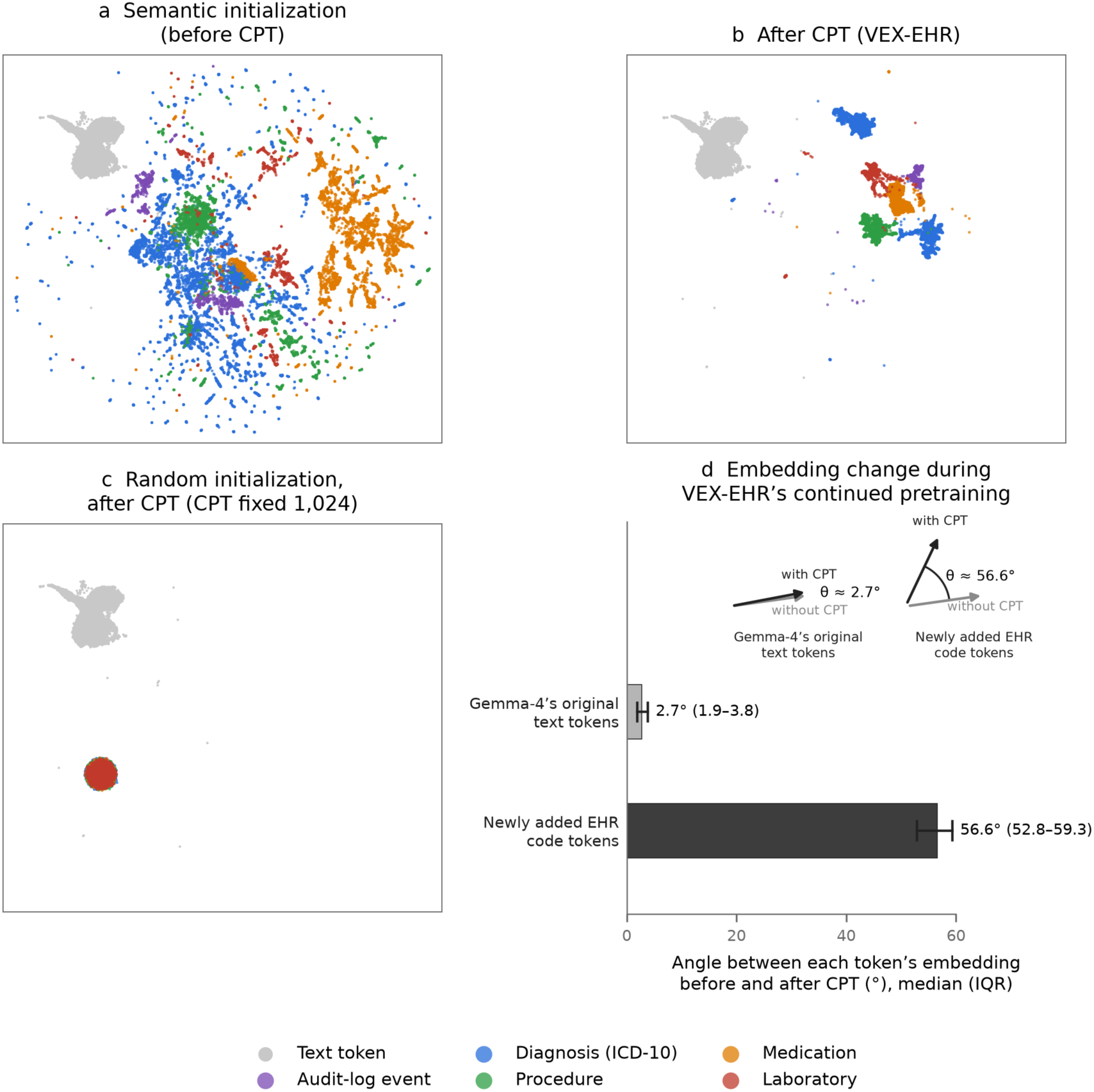
The vocabulary of VEX-EHR in two dimensions: the 20,000 most frequent English text tokens of the backbone (grey) and the 17,601 added clinical and audit-log code tokens (colored by type), in one UMAP projection fitted jointly on every state shown, so positions are comparable across panels. Panels a–c share their axes, so the text tokens cover the same area in each. a, After semantic initialization, before continued pretraining (CPT). b, After CPT (VEX-EHR); diagnoses form two islands associated with the storage order of diagnoses sharing a timestamp. c, Code tokens initialized at random instead of from their descriptions, shown after *CPT fixed 1,024; each of these embeddings turned by a median of 4.3° (interquartile range 3.6–5.2°) during CPT, so they look the same before it. d, Embedding change during the continued pretraining of b (VEX-EHR): the angle between each token’s embedding before and after continued pretraining, median and interquartile range, for Gemma-4’s original text tokens (262,144) and the newly added EHR code tokens (17,601); the insets in the upper right draw this angle, θ, to scale at each group’s median, between a token’s embedding without and with continued pretraining. The other configurations are compared in Supplementary Table S19. Two-dimensional maps are descriptive; the measurements are in Supplementary Section S14*.

Continued pretraining of VEX-EHR moved the code embeddings substantially from their initialization (mean cosine similarity 0.564), whereas the original text-token embeddings barely changed (0.998); it also moved the codes away from the text tokens (mean cosine with the nearest text token, 0.453 to 0.230) and into tighter clusters by type (99.1% same-type neighbors; Figure 5b). At the lower learning rate used for VEX-EHR-Lite and the component analyses, the code embeddings barely moved (cosine similarity to their initialization above 0.99). Randomly initialized codes showed no structure by type, before or after continued pretraining (28.6% and 30.1% same-type neighbors, about the share expected by chance; Figure 5c).

In a training-free probe, we showed the model a code and asked it to pick the code’s own description from 10 candidates of the same category, scoring each candidate by how much more likely it became once the code was shown (chance, 10%; Supplementary Section S15). Right after semantic initialization, before continued pretraining, the model picked the correct description for 26.6% of codes; after continued pretraining, VEX-EHR-Lite picked it for 39.8%, whereas a model whose codes had been randomly initialized picked it for 8.6%.

## Discussion

Adding a health system’s clinical and audit-log codes to a general-purpose LLM as new vocabulary, and continuing its pretraining on that system’s records, produced a model that predicted next-24-hour discharge, 30-day readmission and ICU AKI onset more accurately than an EHR foundation model trained from scratch (CEHR-BERT), and discharge and readmission more accurately than gradient-boosted trees (LightGBM), from which it did not differ clearly on AKI. It needed few labels and remained accurate in the following year, including in patients it had never seen. In a previous study, general LLMs given records serialized into prompts underperformed locally trained models (discharge AUROC 0.63);^7^ given the codes as native vocabulary instead, a general LLM in this study matched or exceeded both locally trained comparators. This suggests that how coded data are presented to a general LLM matters; our comparisons, however, do not separate the contribution of the LLM’s general pretraining from that of its architecture and size.

Prior work has shown that general LLMs can approach EHR-specific models as frozen encoders of serialized records,^8^ when fine-tuned on serialized events,^9,10^ or when coded events are mapped into a frozen LLM through an adapter with description-derived priors.^19^ In those designs the LLM reads codes as text or, in EHRAdapt, as vectors supplied by an external adapter while the LLM itself stays frozen. Here the LLM itself learns the institution’s code tokens and how they occur in its records, at one token per event, and the component analysis shows that this design depends on continued pretraining, whereas models that read serialized text can draw on the backbone’s language directly.

The component analysis identifies what makes the adaptation work. Continued pretraining on the institution’s own event sequences cannot be skipped: new code tokens start only from their descriptions and must still learn how they occur in the institution’s records, and continued pretraining was the only step whose removal lowered accuracy on every task, by 0.03–0.05 AUROC. Initializing each new token from its description is a gain at almost no cost: it needs only each code’s name, added 0.015–0.036 AUROC on two of the three tasks, helped the model learn the institution’s records and organized the codes by type from the start. Description-based initialization also covers all of the institution’s events, not only standard codes: audit-log actions are local workflow events, but the EHR names each with a short English phrase (for example, “Viewed Discharge Instructions”) that serves as its description. The remaining choices (depth, context length and the pretraining corpus) traded accuracy against cost in task-specific ways (see below), and pretraining loss, the measure of how well a model predicts the next event during continued pretraining, did not indicate which configuration would predict clinical outcomes best. A health system adapting a general LLM should therefore compare candidate configurations on labeled validation data for each clinical task. In the natural-language-processing literature, how to initialize the embeddings of an extended vocabulary is an active question,^22–24^ and there is evidence that the content of input embeddings matters less than architecture.^27^ Our finding is consistent with both: description-based initialization helped on two of three tasks and made code tokens partly readable in English, but unlike continued pretraining it did not help on every task.

The learning rate of continued pretraining mattered in a way that suggests how new tokens are learned. At 2×10^−5^, used for VEX-EHR-Lite and the component analyses, the new code embeddings barely moved during continued pretraining (cosine similarity to their initialization above 0.99), so the model adapted almost entirely through its low-rank adapters. At 3×10^−4^, they moved substantially, away from the original text tokens and into tighter clusters by code type (Figure 5; Supplementary Section S14), and accuracy in the 2019–2024 test set improved on discharge and readmission (in 2025, on readmission and AKI but not discharge). A learning rate suited to adjusting a pretrained model’s existing weights may therefore be too low for newly added tokens, whose embeddings have never been trained. Because the learning rate and the number of epochs changed together, this explanation is plausible but not proven; separate learning rates for the new embeddings and for the adapters may be worth testing. In practice, our results favor a higher learning rate during continued pretraining, the stage in which the new token embeddings are learned (3×10^−4^ for one epoch rather than 2×10^−5^ for two), and a low one during fine-tuning (10^−5^ for the adapters; higher values were worse, Supplementary Section S3).

How the components mattered differed by task, in ways that are informative for deployment. Discharge, which has the largest fine-tuning set (38,290 instances), was predicted as well by 6 of the model’s 48 layers as by all 48, so it appears to rest on signals that a shallow model can extract when supervision is abundant. AKI was the most sensitive to the model: removing continued pretraining or most of the layers each cost about 0.05 AUROC. Its label is defined by a change in creatinine, while the model sees only that a creatinine test was ordered, not its result; it has the fewest patients (562 for fine-tuning and 392 for testing), each contributing many predictions; and its accuracy improved when pretraining used full histories instead of only recent events. It was also the one task on which VEX-EHR did not clearly exceed LightGBM, which counts the same codes; neither model sees creatinine values. Readmission sat between the two. Its ceiling is probably set by information the input does not contain: whether a patient returns depends largely on events after discharge, and the label counts only returns to this health system.

Audit-log events, about nine in ten of all input events, record clinicians’ activity, such as which parts of the chart are opened and when,^5^ which can reflect clinical judgment before it appears in structured data and has improved discharge prediction in earlier work.^2^ The adapted model used them: in our occlusion analysis, viewing or printing discharge instructions raised the predicted probability of discharge, and audit-log events accounted for 31% of the AKI prediction’s sensitivity to deleted events (Supplementary Section S8). The improvement in AKI after their removal in the 1,024-token configuration therefore more likely reflects competition for limited input space than a lack of information in audit-log events; at the 4,096 tokens of VEX-EHR, which held 93–100% of the events in the 24-hour window, there is little such competition, although we did not test whether removing audit-log events would change accuracy at this input length.

Adaptation is more expensive to train than a gradient-boosted tree model or a small EHR foundation model built from scratch: against LightGBM, it added 0.049 AUROC on discharge and 0.039 on readmission but no clear gain on AKI. Its efficiency lies elsewhere. Training a model of this size from scratch compute-optimally would use about 240 billion training tokens,^17^ roughly 50 times what our continued pretraining used, whereas adaptation reused the backbone’s general pretraining and trained 0.54% of its weights (low-rank adapters) plus the embedding table. Representing each event as one token also uses the model’s input efficiently: written out as text, each event would take several word pieces, so the same 4,096-token input would hold only a fraction as many events. Most of the 940 GPU-hours went to a one-time continued-pretraining step shared by all tasks, after which each new task needed 23–153 GPU-hours of fine-tuning. Where cost matters more than the last increment of accuracy, it can be reduced: VEX-EHR-Lite used about a quarter of the compute, and a 6-layer model trained 3.7 times as fast with no loss on discharge.

Needing few labels matters for clinical use. Labeled outcomes are often the scarce resource: many outcomes of interest are uncommon, their labels may require chart review by clinicians, and a new task, a small hospital or a changed outcome definition starts with few examples. VEX-EHR exceeded fully trained CEHR-BERT with 5% (discharge) to 10% (readmission) of the labels and fully trained LightGBM with 10% (discharge) to 50% (readmission), so once continued pretraining has been done, a health system could add a prediction task with fewer labels than these comparators need. Adaptation at this cost, most of it for the shared continued pretraining, is within reach of an academic medical center or a shared regional resource, but not of every hospital; sharing adapted models across sites, discussed below, could lower this barrier.

Accuracy was lower in older patients and in men, most clearly for readmission (AUROC 0.650 in patients aged 65 or older against 0.763 in those younger than 45, and 0.669 in men against 0.738 in women) and, by age, for AKI. LightGBM and CEHR-BERT showed gaps in the same direction on the same test instances, VEX-EHR’s gaps were nowhere significantly larger than theirs, and VEX-EHR had the highest AUROC of the three models in every group (Supplementary Section S13). The gaps therefore more likely reflect how hard these outcomes are to predict in these groups, such as older patients with more comorbidity and more complex care, than a property of the adaptation. Before deployment, performance should still be monitored by age and sex, and recalibration or decision thresholds set separately for groups in which the model is less accurate; reweighting or oversampling such groups during fine-tuning is another option, which we did not test.

This study has limitations. It was conducted at a single center. Because the trained weights cannot be shared, we tested the model in the following year, including patients absent from all earlier data; other institutions can repeat the pipeline on their own records, with or without audit-log events. Continued pretraining was run once; the component contrasts were run at one seed in the lower-cost 1,024-token reference configuration and indicate the direction and relative size of each contribution, whose exact magnitude at the main model’s settings may differ; and the full-label discharge model was fine-tuned at one seed because of its cost. We studied one backbone; smaller general models may shift the cost trade-off, and a frozen general LLM with only a classifier trained on its representations, a cheaper design, was not tested. VEX-EHR and CEHR-BERT differ at once in architecture, size and prior pretraining, and we did not continue-pretrain a same-size model from random weights, so the share of the benefit that comes from knowledge acquired in general pretraining is not isolated; description-based initialization of the new tokens, which draws only on that knowledge, added 0.015–0.036 AUROC on two of the three tasks. Laboratory result values were not part of VEX-EHR’s input, and in deployment, events whose codes are outside the 17,601-code vocabulary are omitted from the input; new codes could be added as description-initialized tokens, but this was not tested. Predicted probabilities were too high before recalibration, and decision thresholds did not transfer between fine-tuning runs, so both would need to be set locally before use. Race was linked from a separate table, ethnicity was not analyzed, and smaller racial groups could not be analyzed on their own.

Some inputs may also reflect what the care team already knows or plans, such as viewing discharge instructions or entering a diagnosis for a suspected complication; the model then surfaces the team’s existing judgment rather than anticipating it, which can still help coordinate care. Such signals were also available to the comparators, which used the same 24-hour windows. Discharge instances containing a discharge-day billing code were excluded, and inputs end at the prediction time. AKI diagnosis codes were in the input of 37% of positive AKI predictions; excluding these, VEX-EHR’s AUROC was 0.830–0.848 and LightGBM’s 0.804, a difference similar to that in all AKI predictions. Whether the predictions give earlier warning than clinicians requires prospective evaluation.

Adapting the model to event sequences alone may also have reduced its general language ability, a form of catastrophic forgetting;^28,29^ the text-token embeddings barely changed, but every layer was adapted through low-rank adapters, and we did not measure language-modeling performance. This matters for inputs that combine codes with text, such as laboratory values or notes; mixed-domain continued pretraining, which interleaves event sequences with general or clinical text, is a likely remedy,^30,31^ at a higher cost.

The design also suggests how the approach could scale beyond one institution. Because each new token is defined by its description, another health system’s codes could be added in the same way: standard codes for diagnoses, medications, procedures and laboratory tests share descriptions across institutions, and Epic audit-log actions are named phrases that can be matched across Epic sites. A second institution could therefore adapt the same open-weight backbone to its own codes, adding them as description-initialized tokens and continuing pretraining on its own records, a form of transfer learning that has improved EHR foundation models across sites;^32^ our label-scaling results suggest that it might then need few local labels. Because the backbone’s text vocabulary is retained, laboratory values, note text or retrieved clinical knowledge could also be written into the input beside the codes, supplying, for example, the creatinine values that AKI prediction lacks here. Sharing an adapted model across sites would raise privacy questions, because language models can memorize training data,^33^ and would need corresponding safeguards and data-use agreements. None of these uses was tested. If they hold, health systems could build on general-purpose LLMs without developing each EHR model from scratch.

## Methods

### Data source and cohort

We used de-identified inpatient records from Vanderbilt University Medical Center (VUMC) in the Medical Event Data Standard format (Figure 1): records from 2019–2024 for model development, and records from 2025 for temporal validation only (see Evaluation and statistical analysis). Each patient’s history is one sequence of coded events in time order, drawn from two sources. Clinical events are diagnoses (ICD-10), medications, procedures, laboratory orders with their units, demographics and encounter boundaries. Audit-log events are Epic access actions, the record of which part of the chart a clinician opened or acted on, and when.^2,5^ The vocabulary holds 17,601 codes (Supplementary Table S1). Audit-log actions make up 7% of the codes but 89.9% of event occurrences in the full-history pretraining corpus (93.1% in the corpus of each record’s most recent 1,024 events; Supplementary Table S1). Laboratory result values are not represented.

Patients were split at the patient level into pretraining (170,483 patients), fine-tuning (5,383) and test (3,590) sets; the split is fixed for all models. The records were extracted separately for each calendar year, so a patient admitted in several calendar years contributes one record per year; the pretraining set therefore holds 209,764 records. These are stored year by year from 2019 to 2024: the first 188,787 were used for continued pretraining, and the last 20,977, all from 2024, were held out to monitor the pretraining loss. No fine-tuning or test patient appears in any pretraining corpus.

### Prediction tasks

We studied three inpatient tasks that differ in time horizon, label source and data volume (Table 1).

#### Next-24-hour discharge

For inpatients with a stay of at least 24 hours, a prediction is made at 14:00 on each hospital day from the second day onward. The label is discharge within the next 24 hours. Instances whose input contains a discharge-day billing code (CPT 99238/99239) are excluded, because that code nearly determines the label.

#### 30-day readmission

One prediction is made at discharge. The label is readmission to any VUMC inpatient facility within 30 days.

#### Intensive care unit (ICU) acute kidney injury (AKI) onset

From 6 hours after ICU admission, a prediction is made every 6 hours. The label is KDIGO-defined AKI within 48 hours: a creatinine rise of ≥0.3 mg/dL over the 48-hour minimum, or a value ≥1.5× the 365-day median.^4^ Prediction times at or after a patient’s first AKI in the ICU stay were excluded, as were those with less than 48 hours of ICU follow-up and no AKI.

For every task, the input is the patient’s events in the 24 hours before the prediction time. Table 1 gives the instance counts, patient counts and the share of test instances with the outcome. Because one patient can contribute several prediction instances, the statistical analysis accounts for this clustering (see Evaluation and statistical analysis).

### Adaptation pipeline (VEX-EHR)

The adaptation has four steps (Figure 1): vocabulary extension, semantic (description-based) initialization of the new tokens, continued pretraining and task fine-tuning. The adapted model, VEX-EHR (vocabulary-extended large language model for EHR), is built on Gemma-4 12B. Configuration details are in Supplementary Section S2.

#### Backbone

Gemma-4 12B (12.04 billion parameters, 48 decoder layers, hidden size 3,840; pinned model revision in Supplementary Table S2), text decoder only.^34^

#### Vocabulary extension

The 17,601 clinical and audit-log codes were appended to the backbone’s 262,144-token vocabulary as new tokens (279,745 in total), so each coded event is represented by a single token; writing the event as text would require several subword tokens. The input and output embeddings are tied.

#### Semantic initialization

Each new token’s embedding was set to the mean of the backbone’s own embeddings of the subword tokens of the code’s English description (up to 128 subwords), as in Foresight 2.^25^ Clinical codes use their standard concept name, with the unit for laboratory codes. Audit-log codes use the Epic action name, which is already an English phrase. The new embeddings were rescaled so that their mean norm equals that of the existing vocabulary. Thirty-eight laboratory codes had no description and were initialized from their identifier string.

#### Continued pretraining

Because one institution’s records are far smaller than the corpora on which models of this size are pretrained (see Introduction), we continued pretraining the existing weights. The backbone was trained by next-token prediction on 2,048-token windows at a stride of 1,024 over the full history of each of the 188,787 training records (2,501,553 windows, 4.74 billion tokens), for one epoch (78,173 optimizer steps, effective batch 32). Low-rank adapters (LoRA; rank 16, α = 32, dropout 0.05, on all attention and feed-forward projections; 65.6 million parameters, 0.54% of the model) and the full embedding table were trained.^35^ All other weights were frozen. We used AdamW with a learning rate of 3×10^−4^ (raised from 2×10^−5^, used for VEX-EHR-Lite, at which the new code embeddings barely moved; the two are compared in Supplementary Section S6), cosine decay without warmup, weight decay 0.01 and gradient clipping at 1.0.^36^ Loss on the held-out 2024 records fell from 1.780 after a third of the epoch to 1.718 at its end.

#### Fine-tuning

For each task, the events of the 24-hour input window (up to 4,096; when there were more, the most recent 4,096) were mean-pooled over the final-layer states and passed to a residual multilayer perceptron (MLP) head (two residual blocks; 1.2 million parameters). Training used class-weighted binary cross-entropy in two phases. In phase 1 (3 epochs) only the head was trained, at a learning rate of 10^−3^. In phase 2 LoRA joined, at 10^−5^, for up to 7 more epochs with a patience of 4. The LoRA learning rate was chosen on discharge validation AUROC from 3×10^−6^, 10^−5^, 3×10^−5^ and 10^−4^ with a cheaper configuration (Supplementary Section S3) and kept unchanged. The embedding table stayed frozen, and the effective batch size was 32. A patient-level validation split (10% of the fine-tuning pool) selected the epoch with the highest validation AUROC among phase-2 epochs, ensuring that every selected checkpoint had undergone task-specific LoRA fine-tuning (Supplementary Section S2).

#### VEX-EHR and VEX-EHR-Lite

VEX-EHR, the main model, is continued-pretrained on windows over each record’s full history and fine-tuned at a 4,096-token context. A lower-cost configuration, VEX-EHR-Lite, reads only the most recent 2,048 events of each record, in continued pretraining (two epochs at a learning rate of 2×10^−5^) and in fine-tuning; it is reported alongside VEX-EHR, together with the steps between the two (Supplementary Section S6).

### Comparator models

We compared VEX-EHR with two established approaches trained on the same fine-tuning sets and evaluated on the same test instances (Supplementary Section S4). LightGBM, a gradient-boosted decision-tree model,^37^ was trained on the counts of each of the 17,601 vocabulary codes (clinical and audit-log) in the 24 hours before the prediction time, following the count representation of Zhang et al. for predicting next-day discharge from EHR audit logs.^2^ CEHR-BERT, a transformer EHR foundation model that encodes the timing of events,^14^ was pretrained from scratch on the 170,483 pretraining patients with the same vocabulary and fine-tuned for each task. Its input, as for the other models, is the patient’s events in the 24 hours before the prediction time; unlike the other models, it also receives the recorded numeric values of laboratory and medication events (Supplementary Section S4). Their AUROC and AUPRC were computed with the same bootstrap procedure as for VEX-EHR. LightGBM used fixed hyperparameters, with early stopping on validation AUROC (Supplementary Section S4). Instance-level test predictions were stored for both comparators on every task; all comparisons are paired on identical test instances. Both comparators were also applied without retraining to the 2025 cohort.

### Component analyses

To quantify what each component contributes, we used a lower-cost reference configuration: continued pretraining for two epochs at a learning rate of 2×10^−5^ on the most recent 1,024 events of each record, and fine-tuning on a 1,024-token input. Each comparison changed one component of this reference; data, splits, seed and fine-tuning protocol were unchanged. Continued pretraining: with vs without, and two epochs vs one. New-token initialization: semantic (description-based) vs random (unit-norm Gaussian). Architecture: all 48 layers vs the first 6. Continued-pretraining strategy: a 2,048-vs 1,024-token context in both continued pretraining and fine-tuning, which gives VEX-EHR-Lite; then, from VEX-EHR-Lite, pretraining on sliding windows over full histories with fine-tuning at 4,096 tokens; then one epoch at 3×10^−4^ vs two at 2×10^−5^, which gives VEX-EHR. Input content: without vs with audit-log events in the pretraining corpus and the task inputs. The pretraining loss and downstream AUROC of every configuration are in Supplementary Table S5.

### Evaluation and statistical analysis

The primary metric was test AUROC and the secondary metric AUPRC. Both were reported with 95% percentile bootstrap intervals (1,000 resamples of test instances, seed 42). All VEX-EHR configurations score the same test instances, and label vectors were checked to be identical element by element before any paired comparison.

Test instances are clustered within patients: one patient can contribute many predictions, for example one discharge prediction for each day of a long stay. On AKI, 5,226 test instances come from 392 patients, and the label’s intraclass correlation is 0.76. Treating such instances as independent overstates the precision of a comparison. For comparisons between VEX-EHR configurations we therefore used a patient-clustered bootstrap:^38^ each of 2,000 bootstrap samples resamples patients with replacement and keeps all of each drawn patient’s instances, so that the 95% CI reflects variation between patients. The paired AUROC difference was computed on each sample, and the two-sided *p* value is the bootstrap tail probability. Row-level DeLong tests^39^ are given for reference in Supplementary Table S6.

#### Fine-tuning seeds

Fine-tuning involves random choices (initialization of the prediction head, data order and the training/validation split), so it was repeated with seeds 42, 142 and 242 for readmission and AKI; discharge, whose fine-tuning takes about 153 GPU-hours per run at a 4,096-token context, was run at seed 42 only. Continued pretraining was performed once, so the reported variability reflects fine-tuning. A difference was called consistent only if its patient-clustered 95% CI excluded zero in the same direction at every seed. Headline AUROC is the mean across seeds, with its 95% interval over test instances (Table 2); per-seed values are in Supplementary Table S14.

#### Comparison with established models

Paired differences from LightGBM and CEHR-BERT were estimated with the same patient-clustered bootstrap, in 2019–2024 and in 2025; for readmission and AKI, a difference is called clear only if its interval excludes zero at every fine-tuning seed of VEX-EHR. As a sensitivity analysis for AKI, the trained VEX-EHR (three fine-tuning seeds) and LightGBM models were re-evaluated, without retraining, on the test instances whose 24-hour input contained no AKI diagnosis code (ICD-10 N17; Supplementary Table S21).

#### Temporal validation

A 2025 cohort was built with the same cohort definitions and labels (256,652 discharge, 55,824 readmission and 35,601 AKI instances) and scored by every VEX-EHR model without retraining. We report all instances and two subsets: *new* patients, absent from every 2019–2024 split, and *returning* patients. New and returning patients, being different patients, were compared with an unpaired bootstrap that resampled patients within each group (2,000 draws; for readmission and AKI, the difference was averaged over the three fine-tuning seeds). Continued pretraining used only 2019–2024 records.

#### Fewer labels

With the main model’s continued-pretrained weights fixed, we fine-tuned on fixed patient subsets of 1%, 5%, 10% and 50% of the fine-tuning pool with the main model’s protocol, at seeds 42, 142 and 242 (50%: seed 42 only), and compared the results with LightGBM and, at the same seeds, CEHR-BERT trained on the same patient subsets. AKI at 1% (4 patients) was not estimable, and readmission at 1% is reported descriptively.

#### Calibration, patient groups and cost

Threshold-dependent metrics at a threshold of 0.5 are in Supplementary Table S16. Calibration was assessed before and after a logistic recalibration fitted on each fine-tuning run’s validation predictions (Supplementary Section S12). Decision thresholds were set at the maximum of sensitivity plus specificity (Youden’s index) on each run’s validation predictions (Supplementary Section S13). Discrimination was examined by sex, by age (<45, 45–64 and ≥65 years) and by race, using the groups of a previous study: White and non-White, with unknown and declined race counted as non-White.^7^ For race, the primary measure was VEX-EHR’s AUROC gap between non-White and White patients, judged by the seed rule above (Supplementary Section S13). Compute cost is wall-clock time multiplied by the number of GPUs used, on NVIDIA H100 80 GB GPUs.

### Analyses of the adapted model

Three further analyses describe what the adapted model uses and learns. Occlusion: for a sample of VEX-EHR-Lite’s test instances (500 for discharge, 500 for readmission and 366 for AKI, at most one per patient per class), the events of one category, such as audit-log events or laboratory orders, were deleted from the input and the change in the fine-tuned head’s logit was recorded; a category’s importance share is its mean absolute change divided by the sum over categories, and each deletion was matched by three random deletions of the same number of events (Supplementary Section S8). Vocabulary map: the embeddings of the 17,601 codes and of 20,000 frequent English text tokens were projected jointly to two dimensions with UMAP for display (Figure 5), while all distances were measured in the original 3,840-dimensional space: the cosine similarity of each code embedding to its initialization and to its nearest text token, and the share of each code’s 10 nearest codes that are of its own type (Supplementary Section S14). Probe: without any training, the model was shown each of 500 codes (100 per category) with 10 candidate descriptions of the same category, the code’s own and nine others, in the prompt “Medical code: <code> Meaning: <candidate>”, and each candidate was scored by how much more likely its tokens became once the code was shown (chance, 10%; Supplementary Section S15).

Reporting follows the TRIPOD+AI guideline; the completed checklist is provided as a separate supplementary file.^40^

### Ethics

This study was approved by the Vanderbilt University Medical Center Institutional Review Board (IRB #260543) with a waiver of informed consent. Patients and the public were not involved in the design, conduct or reporting of this study.

### Use of artificial intelligence tools

During the preparation of this work, the authors used Claude (Anthropic) to assist with language editing and code debugging. The authors reviewed all code, checked every reported number against the stored model outputs, and take full responsibility for the content.

## Data Availability

The patient-level data are from Vanderbilt University Medical Center and cannot be shared publicly because they contain protected health information. Access requests may be directed to the corresponding author, subject to institutional approval and a data use agreement. The backbone model (Gemma-4 12B) is publicly available under its published license.

https://github.com/OHPENLab/DischargeWatch

## Code availability

Training and evaluation code, including the statistical analysis, will be made available at https://github.com/OHPENLab/DischargeWatch upon publication and is available to the editors and reviewers on request during peer review; model weights and patient data will not be shared.

## Author contributions

All authors contributed substantially to the work. J.Z. and X.Z. contributed equally to this work. J.Z., X.Z. and Y.C. conceived and designed the study and interpreted the results. J.Z. developed the adaptation method and software, trained the models, performed the experiments and analyses, and drafted and revised the manuscript. X.Z. curated the data, including cohort construction, event extraction and outcome labeling. C.Y. contributed to the interpretation of the results. J.Z.Y. provided some ideas and thoughts on the study. Y.C. provided scientific leadership, supervised and coordinated the project, directed data acquisition and analysis, and acquired funding. X.Z., C.Y., M.B.P., L.L.N., A.N.K., A.W., B.A.M. and Y.C. critically revised the manuscript for important intellectual content. All authors approved the final version and agreed to be accountable for all aspects of the work.

