## Supplementary Information for "Continually pretrained open-weight language models can outperform traditional machine learning in EHR-based clinical prediction"

### S1. Vocabulary and corpus

***Table S1.*** *The 17,601-code vocabulary added to the backbone, by category, and each category's share of token occurrences in the 1,024-token pretraining corpus (the most recent 1,024 events of each of the 209,764 records; 213,978,340 tokens).*

| Category | Codes | Share of tokens | Text used for semantic initialization |
|---|---|---|---|
| Diagnoses (ICD-10) | 7,557 | 0.16% | concept name |
| Medications | 4,513 | 2.32% | concept name |
| Procedures | 2,873 | 0.18%[a] | concept name |
| Demographics and encounter markers | 5 | | concept name |
| Laboratory orders (with unit) | 1,398 | 4.27% | concept name + unit[b] |
| Epic audit-log actions | 1,251 | 93.06% | action name (an English phrase) |
| Special tokens | 4 | — | — |
| Total | 17,601 | 100% | |

[a]Procedures, demographics and encounter markers together. [b]38 laboratory codes have no description anywhere and were initialized from their identifier string. Laboratory result values are not part of the vocabulary; a laboratory token records that a test with that unit was recorded, not its value. Demographic and encounter tokens: 2 sex codes, age at visit, encounter start and encounter end.

The pretraining source concatenates six per-year extractions (2019–2024), so a patient admitted in $k$ calendar years contributes $k$ records: 209,764 records from 170,483 pretraining patients (1.23 per patient). 93.4% of records contain events from a single year. Only 1,550 of 2.77 billion events (0.00006%) carry a 2025 or later timestamp (visit tails), so the corpus respects a 2024 cutoff.

**Pretraining corpora.** The main model, VEX-EHR, is continued-pretrained on 188,787 of the 209,764 records: the source lists records year by year, and the last 20,977, all from 2024, are held out to monitor the pretraining loss. It reads overlapping 2,048-token windows at stride 1,024 over each record's full history: 2.46 billion distinct events and 4.74 billion tokens in its single epoch. VEX-EHR-Lite and the component analyses (Section S6; Table S6) instead read the most recent 2,048 (or 1,024) events of all 209,764 records: 0.43 billion tokens per epoch at 2,048, about one-eleventh as many, with more 2024 records (39,501 against 18,524) but fewer 2024 tokens (81 million against about 543 million). No fine-tuning or test patient is in any pretraining corpus.

### S2. Training configuration

**Model names.** VEX-EHR, the main model, is continued-pretrained on 2,048-token windows over each record's full history (one epoch at a learning rate of $3\times10^{-4}$) and fine-tuned at a 4,096-token context. VEX-EHR-Lite, a lower-cost configuration, reads only the most recent 2,048 events of each record (two epochs at $2\times10^{-5}$) and is fine-tuned at 2,048 tokens. CPT sliding 2,048 at $2\times10^{-5}$ + FT 4,096 (CPT, continued pretraining; FT, fine-tuning) pretrains on VEX-EHR's sliding windows but uses VEX-EHR-Lite's optimizer settings, and is fine-tuned at 4,096 tokens. The reference configuration of the component analyses (Table S6) follows VEX-EHR-Lite's recipe at a 1,024-token context.

***Table S2.*** *Configuration of VEX-EHR (main model; VEX-EHR-Lite differs as given in Section S6) and the training cost of both models in H100 GPU-hours (wall-clock hours multiplied by the number of GPUs, one uninterrupted run; totals computed before rounding).*

| | |
|---|---|
| Backbone | `google/gemma-4-12B`, revision `023679ed352de9bb66cc873c9009ce3482585c08`; text decoder, 48 layers, hidden 3,840, tied input/output embeddings |
| Vocabulary | 262,144 + 17,601 = 279,745 tokens |
| Semantic initialization | mean of the backbone's unscaled embedding rows over the description's subword tokens (cap 128); rows rescaled to the mean norm of the existing vocabulary |
| LoRA | rank 16, α 32, dropout 0.05; q, k, v, o, gate, up, down projections; 65,568,768 parameters |
| Continued pretraining | next-token prediction; full-history corpus: 2,048-token windows, stride 1,024, over full histories (2,501,553 windows); 1 epoch = 78,173 steps; batch 32 |
| | AdamW, lr $3\times10^{-4}$, cosine to 0, no warmup, weight decay 0.01, clip 1.0 |
| | trainable: LoRA + full embedding table (1,139,789,568 parameters) |
| | bf16; gradient checkpointing; 729.7 H100 GPU-hours; final loss 1.6664 (corpus unigram entropy 4.3493; held-out loss 1.7183) |
| Fine-tuning | most recent 4,096 events; mean pooling over final-layer states; residual MLP head (2 residual blocks; 1.2 M parameters); class-weighted BCE (weight = negatives/positives in training rows) |
| | phase 1: 3 epochs, head only, lr $10^{-3}$; phase 2: + LoRA at $10^{-5}$; up to 10 epochs, patience 4 (phase 2 only); embedding table frozen |
| | cosine schedule per optimizer step; weight decay 0.01; clip 1.0; batch 32 |
| | validation: 10% of the fine-tuning pool by patient, balanced on rows and positive rows; selection: best validation AUROC among phase-2 epochs |
| Pretraining tokens | VEX-EHR 4.74 billion (1 epoch); VEX-EHR-Lite 0.85 billion (2 epochs) |
| Training cost | VEX-EHR: continued pretraining 729.7; fine-tuning 209.8 (discharge 153.2, readmission 22.6, AKI 34.1); total 939.6 |
| | VEX-EHR-Lite: continued pretraining 114.2; fine-tuning 136.2 (discharge 101.6, readmission and AKI 34.6); total 250.4 |
| Software | PyTorch 2.9.0, transformers 5.17.0, PEFT 0.20.0, accelerate 1.15.0 |
| Hardware | NVIDIA H100 80 GB GPUs |

**Why phase-2-only selection.** The model evaluated is meant to be the adapted LLM, so selection is restricted to the epochs in which LoRA is trained; selecting over all epochs can return a head trained on the frozen backbone. Across the configurations on the most recent events in Table S5 the two rules select the same epoch on every discharge run (10/10) and every readmission run (9/9); they differ on 2 of 9 AKI runs. For the 1,024-token two-epoch reference, the all-epoch rule would have selected a frozen epoch (test AUROC 0.7509) instead of an adapted epoch (0.8144). For the main model the two rules agree on discharge and differ on readmission and AKI, where a frozen-backbone epoch had the higher validation AUROC (0.7162 against 0.7112; 0.9053 against 0.8976).

**Compute accounting.** GPU-hours in Table S2 are wall-clock hours multiplied by the number of GPUs used, on NVIDIA H100 80 GB GPUs, for one complete, uninterrupted run. VEX-EHR: continued pretraining 729.7; fine-tuning 153.2 (discharge), 22.6 (readmission) and 34.1 (AKI), each including the scoring of the test set. For continued pretraining, this is 78,173 steps at the mean logged step time (700.2 GPU-hours) plus checkpoint saves (23.7) and held-out evaluations (5.8). VEX-EHR-Lite used 114.2 (continued pretraining), 101.6 (discharge) and 34.6 (readmission and AKI) GPU-hours. Scoring with VEX-EHR at 4,096 tokens needs one GPU with at least 27 GB of memory.

### S3. Fine-tuning learning rate

The LoRA learning rate of fine-tuning was chosen with VEX-EHR-Lite (Section S6) on discharge, seed 42, with everything else fixed (the continued-pretraining artifact, the training/validation cut, data order, head learning rate $10^{-3}$, 3 + 7 epochs, phase-2-only selection, patience 4). The rule was specified before the runs: choose the highest best-phase-2 validation AUROC; switch from $10^{-5}$ only if another value exceeds it by at least 0.003, and then replicate it at seeds 142 and 242. Test AUROC played no part in the choice and is reported for every grid point. The chosen value, $10^{-5}$, was kept unchanged for the main model.

***Table S3.*** *LoRA learning-rate grid (discharge, seed 42). Validation AUROC per epoch; epochs 1–3 train the head only and are identical across the grid. Test differences are against $10^{-5}$ on identical rows, with patient-clustered 95% intervals (2,000 draws).*

| LoRA lr | 1 | 2 | 3 | 4 | 5 | 6 | 7 | 8 | 9 | 10 | Best (epoch) | Test | Δ test vs $10^{-5}$ |
|---|---|---|---|---|---|---|---|---|---|---|---|---|---|
| $3\times10^{-6}$ | .7657 | .7810 | .7829 | .8875 | **.8940** | .8936 | .8863 | .8629 | .8329 | – | 0.8940 (5) | 0.9026 | +0.0019 (−0.0007 to +0.0046) |
| $10^{-5}$ (VEX-EHR-Lite; kept) | .7657 | .7810 | .7829 | .8878 | **.8938** | .8932 | .8936 | .8772 | .8422 | – | 0.8938 (5) | 0.9006 | – |
| $3\times10^{-5}$ | .7657 | .7810 | .7829 | .8684 | .8803 | .8861 | **.8866** | .8621 | .8339 | .8339 | 0.8866 (7) | 0.8910 | −0.0096 (−0.0139 to −0.0056) |
| $10^{-4}$ | .7657 | .7810 | .7829 | .5063 | .5152 | .5157 | .5227 | **.5334** | .5092 | .5008 | 0.5334 (8) | 0.5100 | −0.3906 (−0.4030 to −0.3782) |

$3\times10^{-6}$ exceeds $10^{-5}$ on validation by 0.0002, below the 0.003 threshold, so $10^{-5}$ is kept; on test the two do not differ. At $10^{-4}$ training diverged and validation AUROC stayed near 0.5. A dash marks an epoch not run after early stopping (patience 4).

### S4. Comparator models

**LightGBM.** Gradient-boosted decision trees (main-text reference 37) were trained for each task on the counts of each of the 17,601 vocabulary codes (clinical and audit-log) in the 24 hours before the prediction time, following the count representation of Zhang et al. for next-day discharge prediction from EHR audit logs (main-text reference 2). Hyperparameters were fixed and the same for all three tasks (31 leaves, maximum depth 15, learning rate 0.1, feature fraction 0.75, bagging fraction 0.77, at least 7 instances per leaf, class weighting for the imbalanced labels), with up to 981 trees and early stopping after 100 rounds without improvement in AUROC on a 10% validation split of the fine-tuning set (stratified by instance for discharge and readmission and by patient for AKI). LightGBM was trained on the same fine-tuning sets and, in the label-scaling experiments, on the same patient subsets as VEX-EHR (Section S10), and was evaluated on the same test instances.

**CEHR-BERT.** CEHR-BERT (main-text reference 14) is a bidirectional transformer that represents a patient's coded history together with the timing of events. It was pretrained from scratch on the 170,483 pretraining patients by masked-token prediction, with the same 17,601-code vocabulary as VEX-EHR, audit-log codes included, plus its own visit and time-interval tokens. It was fine-tuned for each task for 10 epochs, keeping the epoch with the highest validation AUROC. Its input for each prediction is the patient's events in the 24 hours before the prediction time, the same window as for VEX-EHR and LightGBM, grouped into visits by calendar day as CEHR-BERT requires. Its input also carries the recorded numeric values of laboratory and medication events; VEX-EHR and LightGBM do not receive these values. With all labels, it was fine-tuned once (seed 42); in the label-scaling experiments, also at seeds 142 and 242 (Section S10).

**Evaluation.** Test AUROC and AUPRC of both comparators, with 95% percentile bootstrap intervals over test instances (1,000 resamples), were computed with the same procedure as for VEX-EHR. Instance-level test predictions were stored for both comparators on every task; differences from VEX-EHR are paired (Table S4), with the patient-clustered bootstrap of main-text Methods. Both comparators were also applied, without retraining, to the 2025 cohort and paired with VEX-EHR in the same way (Table S11). LightGBM's stored predictions in the label-scaling experiments were paired with VEX-EHR's in the same way (Section S10).

***Table S4.*** *VEX-EHR against the comparators on identical 2019–2024 test instances: test AUROC, the paired AUROC difference with its patient-clustered bootstrap 95% interval (2,000 draws), row-level DeLong p and the clustered bootstrap p (<0.001 means no draw crossed zero). LightGBM: all tasks (early stopping on validation AUROC; AUROCs as in main-text Table 2). CEHR-BERT: all tasks, fine-tuned once (seed 42). VEX-EHR: discharge seed 42 (26,300 instances); readmission and AKI seeds 42, 142 and 242.*

| Comparator | Task | Seed | AUROC, VEX-EHR | AUROC, comparator | ΔAUROC (95% CI) | DeLong p | Clustered p |
|---|---|---|---|---|---|---|---|
| LightGBM | Discharge | 42 | 0.9172 | 0.8680 | +0.0491 (+0.0429 to +0.0556) | $1\times10^{-67}$ | <0.001 |
| | Readmission | 42 | 0.7090 | 0.6767 | +0.0323 (+0.0128 to +0.0520) | 0.0005 | <0.001 |
| | | 142 | 0.7195 | 0.6767 | +0.0428 (+0.0219 to +0.0641) | $7\times10^{-6}$ | <0.001 |
| | | 242 | 0.7174 | 0.6767 | +0.0406 (+0.0222 to +0.0609) | $1\times10^{-5}$ | <0.001 |
| | AKI | 42 | 0.8424 | 0.8282 | +0.0143 (−0.0188 to +0.0445) | 0.06 | 0.348 |
| | | 142 | 0.8412 | 0.8282 | +0.0131 (−0.0201 to +0.0442) | 0.1 | 0.397 |
| | | 242 | 0.8588 | 0.8282 | +0.0306 (−0.0033 to +0.0639) | $5\times10^{-5}$ | 0.072 |
| CEHR-BERT | Discharge | 42 | 0.9172 | 0.8480 | +0.0692 (+0.0617 to +0.0766) | $5\times10^{-92}$ | <0.001 |
| | Readmission | 42 | 0.7090 | 0.6501 | +0.0589 (+0.0346 to +0.0850) | $1\times10^{-6}$ | <0.001 |
| | | 142 | 0.7195 | 0.6501 | +0.0693 (+0.0439 to +0.0971) | $3\times10^{-8}$ | <0.001 |
| | | 242 | 0.7174 | 0.6501 | +0.0672 (+0.0439 to +0.0923) | $1\times10^{-8}$ | <0.001 |
| | AKI | 42 | 0.8424 | 0.7914 | +0.0510 (+0.0122 to +0.0875) | $1\times10^{-7}$ | 0.009 |
| | | 142 | 0.8412 | 0.7914 | +0.0498 (+0.0165 to +0.0825) | $5\times10^{-7}$ | 0.003 |
| | | 242 | 0.8588 | 0.7914 | +0.0674 (+0.0420 to +0.0965) | $4\times10^{-14}$ | <0.001 |

### S5. All configurations and component contributions

***Table S5.*** *Every configuration trained, with its continued-pretraining loss and test AUROC. Loss = mean negative log-likelihood of the next token during continued pretraining (nats per token; lower means the next event is predicted better). Gap = corpus unigram entropy − final training loss (nats per token), the margin by which context beats a frequency table; raw losses are comparable only within one corpus. Run-to-run variation in loss is about 0.015–0.018 nats, so Gap differences below about 0.03 are not interpretable. Seed 42. The main model is in the first block.*

| Configuration | Loss | Unigram | Gap | Discharge | Readmission | AKI |
|---|---|---|---|---|---|---|
| *4,096-token context, full-history corpus* | | | | | | |
| Main model ($3\times10^{-4}$, one epoch) | 1.6664 | 4.3493 | 2.6829 | 0.9172 | 0.7090 | 0.8424 |
| CPT sliding 2,048 at $2\times10^{-5}$ + FT 4,096 (two epochs) | 1.7923 | 4.3493 | 2.5570 | 0.9091 | 0.6903 | 0.8556 |
| *2,048-token context* | | | | | | |
| VEX-EHR-Lite | 1.8593 | 4.1736 | 2.3143 | 0.9006 | 0.7090 | 0.8078 |
| Sampled-window corpus[a] | 1.9398 | 4.3091 | 2.3693 | 0.8994 | 0.7009 | 0.8151 |
| *1,024-token context* | | | | | | |
| Reference (2 epochs) | 1.8855 | 4.1359 | 2.2504 | 0.8822 | 0.6892 | 0.8144 |
| One pretraining epoch | 1.9484 | 4.1359 | 2.1875 | 0.8859 | 0.6913 | 0.7904 |
| No continued pretraining | — | — | — | 0.8311 | 0.6584 | 0.7610 |
| Random initialization | 1.9534 | 4.1359 | 2.1825 | 0.8668 | 0.6529 | 0.8038 |
| First 6 of 48 layers | 2.3395 | 4.1359 | 1.7964 | 0.8813 | 0.6754 | 0.7614 |
| Layers 0–2 and 45–47[b] | 2.1928 | 4.1359 | 1.9431 | 0.8725 | 0.6781 | 0.7682 |
| Sampled-window corpus[a] | 1.9965 | 4.3254 | 2.3289 | 0.8916 | 0.6955 | 0.8183 |
| Audit-log events removed[c] | 1.4655 | 5.4071 | 3.9416 | 0.8867 | 0.6959 | 0.8491 |

[a]One window per record at a uniformly sampled position instead of the most recent events; the token budget is identical. [b]A second 6-layer arrangement (first three and last three layers). [c]Removing audit events changes the corpus, so its loss and Gap are on a different scale. Pretraining loss does not order configurations by downstream result: the 6-layer model has the smallest Gap and ties the reference on discharge, and the largest Gap among audit-bearing corpora of recent events (sampled windows, 2,048) is best on no task. The full-history corpus has its own unigram entropy (4.3493) and is not comparable with the others.

***Table S6.*** *Contribution of each adaptation component (part A: main-text Figure 3). Each row gives the test AUROC of configuration A minus that of configuration B (A vs B), paired, with a patient-clustered bootstrap 95% interval (2,000 draws), the row-level DeLong p, the patient-clustered bootstrap p (two-sided tail probability; <0.001 means no draw crossed zero) and the ratio of the clustered to the DeLong standard error. Part A changes one component of the reference configuration at a time and repeats continued pretraining and fine-tuning; part B gives the steps from VEX-EHR-Lite to VEX-EHR (the two models themselves: main-text Table 2). CPT, continued pretraining; FT, fine-tuning. CPT fixed n uses the most recent n events of each record; CPT sliding n uses n-token windows sliding over each record; FT n uses the most recent n tokens of the 24-hour input. The reference also uses description-based initialization, 48 layers and audit-log events. Seed 42 throughout.*

| Comparison (A vs B) | Task | ΔAUROC (95% CI) | DeLong p | Clustered p | SE ratio |
|---|---|---|---|---|---|
| *A. One component changed from the reference configuration (CPT fixed 1,024, two epochs at $2\times10^{-5}$; FT 1,024; AUROC 0.8822 / 0.6892 / 0.8144)* | | | | | |
| CPT: With vs without CPT | Discharge | +0.051 (+0.044, +0.057) | $7\times10^{-63}$ | <0.001 | 1.08 |
| | Readmission | +0.031 (+0.013, +0.048) | 0.0002 | <0.001 | 1.08 |
| | AKI | +0.053 (+0.026, +0.083) | $2\times10^{-10}$ | <0.001 | 1.74 |
| Initialization: Description-based vs random | Discharge | +0.015 (+0.010, +0.021) | $2\times10^{-8}$ | <0.001 | 1.03 |
| | Readmission | +0.036 (+0.020, +0.055) | $1\times10^{-5}$ | <0.001 | 1.10 |
| | AKI | +0.011 (−0.016, +0.037) | 0.14 | 0.41 | 1.87 |
| Transformer depth: 48 vs 6 layers | Discharge | +0.001 (−0.004, +0.006) | 0.71 | 0.74 | 1.02 |
| | Readmission | +0.014 (+0.001, +0.028) | 0.030 | 0.041 | 1.08 |
| | AKI | +0.053 (+0.025, +0.083) | $6\times10^{-14}$ | <0.001 | 2.10 |
| CPT epochs: Two vs one epoch | Discharge | −0.004 (−0.008, +0.000) | 0.047 | 0.062 | 1.05 |
| | Readmission | −0.002 (−0.013, +0.009) | 0.69 | 0.76 | 1.09 |
| | AKI | +0.024 (+0.006, +0.046) | $9\times10^{-10}$ | 0.010 | 2.65 |
| Context length: CPT fixed 2,048 + FT 2,048 (VEX-EHR-Lite) vs CPT fixed 1,024 + FT 1,024 | Discharge | +0.018 (+0.014, +0.023) | $1\times10^{-15}$ | <0.001 | 1.05 |
| | Readmission | +0.020 (+0.006, +0.034) | 0.003 | 0.005 | 1.08 |
| | AKI | −0.007 (−0.030, +0.016) | 0.28 | 0.56 | 1.97 |
| Input content: Without vs with audit-log events | Discharge | +0.005 (−0.001, +0.010) | 0.095 | 0.12 | 1.05 |
| | Readmission | +0.007 (−0.009, +0.022) | 0.42 | 0.45 | 1.01 |
| | AKI | +0.035 (+0.005, +0.065) | $3\times10^{-6}$ | 0.023 | 2.09 |

| Comparison (A vs B) | Task | ΔAUROC (95% CI) | DeLong p | Clustered p | SE ratio |
|---|---|---|---|---|---|
| *B. From VEX-EHR-Lite to VEX-EHR (CPT fixed 2,048, two epochs at 2×10$^{-5}$; FT 2,048; AUROC 0.9006 / 0.7090 / 0.8078)* | | | | | |
| CPT window: CPT sliding 2,048 + FT 4,096 vs CPT fixed 2,048 + FT 2,048 (VEX-EHR-Lite) | Discharge | +0.008 (+0.005, +0.012) | 2×10$^{-5}$ | <0.001 | 0.99 |
| | Readmission | −0.019 (−0.035, −0.001) | 0.037 | 0.043 | 0.99 |
| | AKI | +0.048 (+0.018, +0.078) | 1×10$^{-12}$ | 0.002 | 2.21 |
| CPT learning rate: 3×10$^{-4}$, one epoch vs 2×10$^{-5}$, two epochs (both CPT sliding 2,048 + FT 4,096; the first is VEX-EHR) | Discharge | +0.008 (+0.004, +0.012) | 3×10$^{-5}$ | <0.001 | 1.05 |
| | Readmission | +0.019 (+0.000, +0.037) | 0.039 | 0.047 | 1.01 |
| | AKI | −0.013 (−0.047, +0.019) | 0.033 | 0.42 | 2.67 |
| Overall: VEX-EHR vs VEX-EHR-Lite | Discharge | +0.017 (+0.012, +0.021) | 1×10$^{-14}$ | <0.001 | 1.08 |
| | Readmission | +0.000 (−0.012, +0.013) | 1.00 | 0.996 | 1.02 |
| | AKI | +0.035 (+0.003, +0.066) | 1×10$^{-6}$ | 0.027 | 2.25 |

"Without audit-log events" removes them from the pretraining corpus before truncation and from every task input. In part B, the learning rate and the number of epochs change together, and the Overall row is a direct paired comparison (Section S6); the pretraining loss of every configuration is in Table S5. On discharge and readmission patient clustering changes the standard error by a factor of 0.99–1.10; on AKI it widens it by 1.7–2.7, because each AKI patient contributes 13.3 test instances on average and the label's intraclass correlation is 0.76. Conclusions on AKI should be drawn from the clustered column only.

### S6. Continued-pretraining strategy: from VEX-EHR-Lite to VEX-EHR

VEX-EHR-Lite read the most recent 2,048 events of each of the 209,764 records: continued pretraining for two epochs (13,110 optimizer steps at effective batch 32) at a learning rate of 2×10$^{-5}$, and fine-tuning at 2,048 tokens, with every other setting of the main model. Because the most recent 2,048 events cover a small part of a long record, the main model, VEX-EHR, is continued-pretrained on the full-history corpus, 2,048-token sliding windows with stride 1,024 over each training record's full history (2,501,553 windows), with the same vocabulary extension and semantic initialization, at 3×10$^{-4}$ for one epoch (78,173 steps; 730 GPU-hours). CPT sliding 2,048 at 2×10$^{-5}$ + FT 4,096 pretrains on VEX-EHR's corpus but uses VEX-EHR-Lite's optimizer settings (two epochs, 156,348 optimizer steps; about 1,400 H100 GPU-hours, fine-tuning about 200 more). Both were fine-tuned at a 4,096-token context with the main model's protocol, seed 42. Between VEX-EHR-Lite and CPT sliding 2,048 at 2×10$^{-5}$ + FT 4,096, the pretraining corpus and the context change together; between CPT sliding 2,048 at 2×10$^{-5}$ +

FT 4,096 and the main model the learning rate and the epoch count change together; neither pair can be separated.

Two discharge test instances (both negative) had no events in the input window and are excluded; all models are compared on the remaining 26,300 instances.

***Table S7.*** *From VEX-EHR-Lite to VEX-EHR (seed 42): test AUROC on the 2019–2024 test instances and in the 2025 cohort, and paired differences with patient-clustered 95% intervals (2,000 draws, seed 42) on the 2019–2024 instances and in the 2025 cohort. For 2025 readmission and AKI, the two models were also compared at fine-tuning seeds 142 and 242, each at the same seed (last rows; discharge was fine-tuned at seed 42 only).*

| | Discharge ($n$ = 26,300) | Readmission ($n$ = 5,471) | AKI ($n$ = 5,226) |
|---|---|---|---|
| *2019–2024 test instances* | | | |
| VEX-EHR-Lite (2,048; 2×10$^{-5}$, two epochs) | 0.9006 | 0.7090 | 0.8078 |
| CPT sliding 2,048 at 2×10$^{-5}$ + FT 4,096 (two epochs) | 0.9091 | 0.6903 | 0.8556 |
| VEX-EHR (4,096; 3×10$^{-4}$, one epoch) | 0.9172 | 0.7090 | 0.8424 |
| CPT sliding 2,048 at 2×10$^{-5}$ + FT 4,096 − VEX-EHR-Lite | +0.0084 (+0.0046, +0.0122) | −0.0187 (−0.0349, −0.0009) | +0.0478 (+0.0185, +0.0778) |
| VEX-EHR − CPT sliding 2,048 at 2×10$^{-5}$ + FT 4,096 | +0.0081 (+0.0042, +0.0122) | +0.0187 (+0.0003, +0.0372) | −0.0132 (−0.0474, +0.0186) |
| VEX-EHR − VEX-EHR-Lite | +0.0165 (+0.0122, +0.0212) | +0.0000 (−0.0121, +0.0125) | +0.0346 (+0.0033, +0.0661) |
| *2025 cohort* | | | |
| VEX-EHR-Lite | 0.974 | 0.687 | 0.760 |
| CPT sliding 2,048 at 2×10$^{-5}$ + FT 4,096 (two epochs) | 0.971 | 0.682 | 0.794 |
| VEX-EHR (4,096; 3×10$^{-4}$, one epoch) | 0.963 | 0.705 | 0.803 |
| CPT sliding 2,048 at 2×10$^{-5}$ + FT 4,096 − VEX-EHR-Lite | −0.0026 (−0.0033, −0.0019) | −0.0047 (−0.0105, +0.0014) | +0.0339 (+0.0249, +0.0431) |
| VEX-EHR − CPT sliding 2,048 at 2×10$^{-5}$ + FT 4,096 | −0.0086 (−0.0098, −0.0073) | +0.0231 (+0.0167, +0.0291) | +0.0092 (+0.0014, +0.0164) |
| VEX-EHR − VEX-EHR-Lite, seed 42 | −0.0112 (−0.0124, −0.0100) | +0.0183 (+0.0140, +0.0227) | +0.0430 (+0.0334, +0.0526) |
| VEX-EHR − VEX-EHR-Lite, seed 142 | — | +0.0031 (−0.0023, +0.0081) | +0.0320 (+0.0215, +0.0424) |
| VEX-EHR − VEX-EHR-Lite, seed 242 | — | +0.0340 (+0.0283, +0.0399) | +0.0415 (+0.0312, +0.0515) |
| VEX-EHR − VEX-EHR-Lite, mean of three seeds | — | +0.0185 | +0.0388 |

Validation AUROC (selected epoch): CPT sliding 2,048 at 2×10$^{-5}$ + FT 4,096, discharge 0.9037 (7), readmission 0.7356 (7), AKI 0.9048 (4); main model 0.9136 (6), 0.7112 (4), 0.8976 (5). VEX-EHR-Lite, three seeds: discharge 0.8990 ± 0.0023, readmission 0.7082 ± 0.0042, AKI 0.8185 ± 0.0093. Against VEX-EHR-Lite, the main model is better on discharge and AKI and equal on readmission; on the 2025 AKI cohort it exceeded VEX-EHR-Lite at every seed (mean difference +0.0388).

**Interpretation.** Continued pretraining on 2,048-token sliding windows over full histories with fine-tuning at 4,096 tokens, instead of the most recent 2,048 events at 2,048 tokens, raised AKI by 0.048 and discharge by 0.008 and lowered readmission by 0.019, at about six times the cost. Raising the learning rate to $3\times10^{-4}$ for one epoch then recovered readmission (+0.019) and added 0.008 on discharge at half the full-history cost, and left AKI level with the previous step (−0.013, interval crossing zero). In 2025 the full-history corpus raised AKI; the higher learning rate raised readmission but lowered discharge relative to both earlier steps, so the main model is not the best of the three on every task in 2025.

### S7. AKI: audit-log crowding and pretraining histories

Figure S1 shows the AKI analyses discussed in the main text: occlusion attribution by event category (Section S8) and the AKI test AUROC of the configurations of Table S6 and Section S6. Table S8 gives, for all three tasks, the clinical events that the 1,024-event inputs of the reference configuration hold with and without audit-log events.

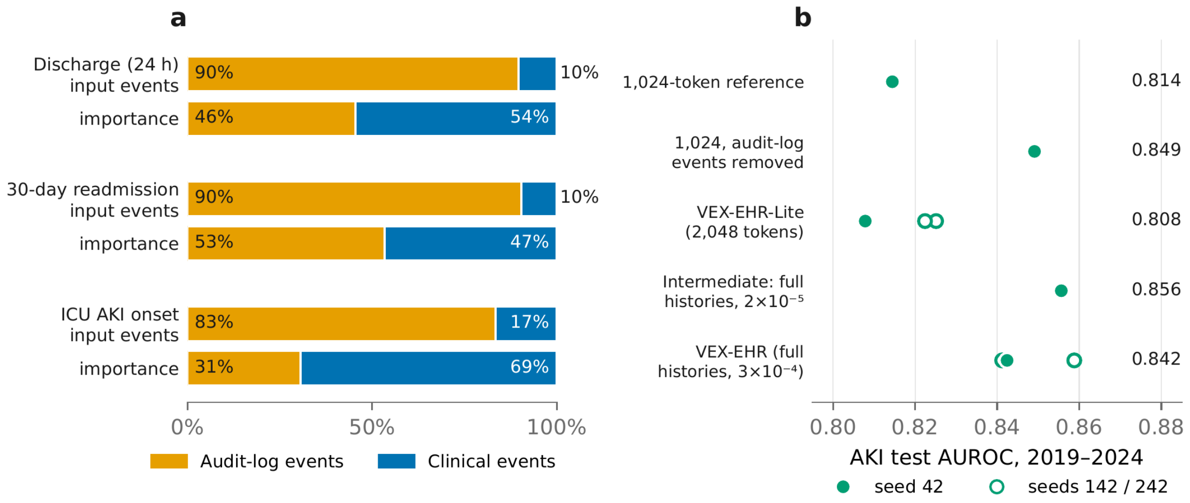

***Figure S1.*** *AKI: audit-log crowding and pretraining histories. a, Occlusion attribution of VEX-EHR-Lite (seed 42; 500, 500 and 366 test instances): for each task, the share of input events that are audit-log versus clinical events, and the share of the prediction's sensitivity to deleting each*

*group. On AKI, clinical events are 17% of the input but carry 69% of the importance. Events of other kinds (<0.3%) are omitted. b, AKI test AUROC (2019–2024) of VEX-EHR configurations: the 1,024-token reference, the same with audit-log events removed from pretraining and inputs, VEX-EHR-Lite (2,048 tokens), the model pretrained on the full-history sliding-window corpus and fine-tuned at 4,096 tokens with VEX-EHR-Lite's optimizer settings, and the main model (the same corpus at a learning rate of $3\times10^{-4}$ for one epoch). Filled markers: seed 42; open markers: seeds 142 and 242. Paired differences are in Tables S6 and S7. Details in Sections S6 and S8.*

***Table S8.*** *Clinical events in the fine-tuning inputs of the 1,024-token reference configuration, with and without audit-log events (2019–2024; all labeled instances, built as for fine-tuning). The input holds the most recent 1,024 events of the 24-hour window. With / without: with audit-log events (the reference) / audit-log events removed from pretraining and inputs (Table S6, input content). Truncated: inputs whose events did not all fit in 1,024 tokens; for AKI, whose fine-tuning cache stores only the most recent 1,024 events, input lengths were taken from a cache of the most recent 8,192 events (0.5% of inputs are longer). Creatinine-related: any of the 15 laboratory codes whose description contains "creatinine".*

| Measure | Discharge (64,592) | Readmission (13,831) | AKI (13,285) |
|---|---|---|---|
| Audit-log share of input events | 88.5% | 91.7% | 84.0% |
| Clinical events per input, median (IQR), with / without | 104 (74–133) / 153 (109–222) | 77 (47–105) / 117 (81–152) | 136 (99–187) / 282 (189–437) |
| Inputs truncated at 1,024 events, with / without | 78.0% / 0.3% | 74.4% / 0.0% | 90.9% / 1.9% |
| No creatinine-related laboratory order in the input, with / without | 40.7% / 30.1% | 57.2% / 43.8% | 55.9% / 22.1% |

### S8. Occlusion attribution

For a test sample of VEX-EHR-Lite (seed 42; occlusion was not repeated for the main model), consisting of 500 discharge and 500 readmission instances (half positive) and the 366 AKI instances that qualified (116 positive), at most one per patient per class, events of one category were deleted from the input and the change in the fine-tuned head's logit was recorded. Importance share is a category's mean absolute change divided by the sum over categories; token share is its share of input events; enrichment is their ratio, with a bootstrap interval over instances. Each deletion was matched by three random deletions of the same number of events. The unmodified input reproduced the stored test probabilities (median absolute difference

0.0005–0.0012). Integrated gradients were not used because their completeness check failed on every instance in a pilot.

***Table S9.*** *Occlusion by event category, VEX-EHR-Lite. Token share / importance share / enrichment (95% CI).*

| Category | Discharge (500) | Readmission (500) | AKI (366) |
|---|---|---|---|
| Audit-log | 0.896 / 0.455 / 0.51 (0.49, 0.53) | 0.904 / 0.534 / 0.59 (0.57, 0.61) | 0.834 / 0.306 / 0.37 (0.34, 0.39) |
| All clinical | 0.104 / 0.544 / 5.23 (4.82, 5.64) | 0.095 / 0.456 / 4.80 (4.34, 5.28) | 0.166 / 0.692 / 4.17 (3.77, 4.65) |
| laboratory orders | 0.069 / 0.075 / 1.08 (0.94, 1.23) | 0.058 / 0.139 / 2.39 (2.17, 2.63) | 0.132 / 0.212 / 1.60 (1.39, 1.86) |
| medications | 0.031 / 0.288 / 9.37 (8.01, 10.66) | 0.033 / 0.249 / 7.47 (6.49, 8.51) | 0.027 / 0.401 / 14.58 (12.95, 16.34) |
| diagnoses (ICD-10) | 0.003 / 0.094 / 34.3 (28.9, 40.4) | 0.003 / 0.051 / 19.5 (16.0, 23.8) | 0.005 / 0.062 / 13.2 (10.9, 15.9) |
| procedures | 0.001 / 0.086 / 77.2 (66.1, 90.2) | 0.001 / 0.017 / 17.4 (14.4, 20.7) | 0.002 / 0.017 / 10.7 (8.7, 13.1) |
| Mean \|Δ\| logit, all clinical vs random | 1.85 vs 0.19 | 0.46 vs 0.05 | 1.51 vs 0.23 |

Other events are below 0.3% of tokens on every task and are omitted. Clinical enrichment is similar in positive and negative instances (discharge 4.71 / 5.70, readmission 5.11 / 4.48, AKI 4.06 / 4.22). Audit-log events act jointly: the summed single-code effects recover only 15–40% of the category effect. Deleting events changes the input distribution; the random deletions control for this only in part. One seed.

**Interpretation.** On AKI, clinical events are 17% of the input but carry 69% of the importance; audit-log events, 83% of the input, carry 31%. This is consistent with the crowding suggested by the audit-free models (Section S7; Table S8), but importance shares alone do not establish it. On discharge, audit-log events are informative in bulk (45% of the importance; individual actions such as viewing the discharge instructions or printing raise the predicted probability) though under-weighted per event. Serum creatinine orders are not among the most influential AKI codes, consistent with laboratory results being absent from the input; the model relies instead on markers of ICU severity (inspired oxygen fraction, blood gases, broad-spectrum antibiotics, sedation).

### S9. Temporal validation (2025)

The 2025 temporal test set was built with the same cohort definitions and labels as the 2019–2024 data and scored by every VEX-EHR model without retraining. Before any 2025 score was used, each of the seven VEX-EHR models (discharge one seed; readmission and AKI three each) re-scored its 2019–2024 test rows and reproduced its stored predictions exactly (maximum absolute difference 0). Labels were checked to be identical element by element. *New*: patients in no 2019–2024 split, including pretraining; *returning*: the rest. The main model was pretrained on 18,524 records of 2024 (46.9%; the rest are held out to monitor the pretraining loss); VEX-EHR-Lite (Section S6) used all 39,501.

***Table S10.*** *2025 temporal validation: test AUROC of VEX-EHR by cohort (readmission and AKI: three fine-tuning seeds; discharge: seed 42).*

| Task | Cohort (instances / patients) | AUROC, mean of seeds | Seed 42 | Seed 142 | Seed 242 |
|---|---|---|---|---|---|
| Discharge | all (256,652 / 40,499) | 0.963 | 0.9627 | — | — |
| | new (159,182 / 27,823) | 0.966 | 0.9661 | — | — |
| | returning (97,470 / 12,676) | 0.955 | 0.9553 | — | — |
| Readmission | all (55,824 / 42,234) | 0.703 | 0.7050 | 0.6972 | 0.7074 |
| | new (35,192 / 29,113) | 0.711 | 0.7129 | 0.7065 | 0.7130 |
| | returning (20,632 / 13,121) | 0.654 | 0.6540 | 0.6488 | 0.6584 |
| AKI | all (35,601 / 3,264) | 0.805 | 0.8034 | 0.7969 | 0.8134 |
| | new (25,849 / 2,315) | 0.795 | 0.7948 | 0.7868 | 0.8045 |
| | returning (9,752 / 949) | 0.821 | 0.8190 | 0.8158 | 0.8293 |

***Table S11.*** *2025, without retraining: VEX-EHR against CEHR-BERT and LightGBM on identical instances (instances / patients). AUROC of each model (VEX-EHR: mean over its fine-tuning seeds) and the paired AUROC difference of VEX-EHR from each comparator with the patient-clustered bootstrap (2,000 draws); for three seeds, the mean difference and, in parentheses, the lowest lower and highest upper 95% bound across seeds. Every interval excludes zero except those of VEX-EHR against LightGBM on AKI.*

| Task | Cohort (instances / patients) | AUROC, VEX-EHR | AUROC, CEHR-BERT | AUROC, LightGBM | ΔAUROC vs CEHR-BERT (95% CI) | ΔAUROC vs LightGBM (95% CI) | VEX-EHR seeds |
|---|---|---|---|---|---|---|---|
| Discharge | all (256,652 / 40,499) | 0.963 | 0.931 | 0.941 | +0.031 (+0.029 to +0.033) | +0.021 (+0.020 to +0.023) | 42 |
| | new (159,182 / 27,823) | 0.966 | 0.932 | 0.945 | +0.034 (+0.032 to +0.036) | +0.021 (+0.020 to +0.023) | 42 |
| | returning (97,470 / 12,676) | 0.955 | 0.930 | 0.934 | +0.025 (+0.022 to +0.029) | +0.021 (+0.018 to +0.024) | 42 |
| Readmission | all (55,824 / 42,234) | 0.703 | 0.611 | 0.665 | +0.092 (+0.078 to +0.104) | +0.039 (+0.027 to +0.048) | 42 / 142 / 242 |
| | new (35,192 / 29,113) | 0.711 | 0.617 | 0.674 | +0.094 (+0.079 to +0.106) | +0.036 (+0.025 to +0.046) | 42 / 142 / 242 |
| | returning (20,632 / 13,121) | 0.654 | 0.573 | 0.624 | +0.081 (+0.064 to +0.097) | +0.030 (+0.015 to +0.043) | 42 / 142 / 242 |
| AKI | all (35,601 / 3,264) | 0.805 | 0.731 | 0.787 | +0.074 (+0.054 to +0.094) | +0.018 (−0.002 to +0.037) | 42 / 142 / 242 |
| | new (25,849 / 2,315) | 0.795 | 0.727 | 0.778 | +0.068 (+0.045 to +0.090) | +0.017 (−0.006 to +0.041) | 42 / 142 / 242 |
| | returning (9,752 / 949) | 0.821 | 0.729 | 0.803 | +0.093 (+0.064 to +0.122) | +0.019 (−0.007 to +0.044) | 42 / 142 / 242 |

Audit-free models on 2025 AKI (1,024-token context, seed 42): an audit-free model scored 0.7880 against 0.7662 for the audit-bearing model at the same context (+0.0218, patient-clustered 95% CI +0.0111 to +0.0322). On the 2025 AKI cohort, the main model exceeded VEX-EHR-Lite at every seed (mean difference +0.0388; Section S6).

### S10. Fine-tuning label scaling

We fine-tuned the main model's continued-pretrained weights on fixed patient subsets of 1%, 5%, 10% and 50% of the fine-tuning pool, with the main model's protocol unchanged, at seeds 42, 142 and 242 (50%: seed 42 only). LightGBM was trained on the same patient subsets and evaluated on the same test instances (Section S4); differences from it were tested in pairs with the patient-clustered bootstrap and the seed rule of main-text Methods (50%: seed 42 only, judged on that seed). CEHR-BERT was fine-tuned on the same subsets at seeds 42, 142 and 242

(50%: seed 42 only) and is reported descriptively; at several small fractions its predictions were nearly constant (Table S12). Readmission at 1% is descriptive because the models are close to chance; AKI at 1% (4 patients) is not estimable.

***Table S12.*** *Label scaling: test AUROC of VEX-EHR (seeds 42 / 142 / 242 and their mean) and of CEHR-BERT (seeds 42 / 142 / 242 at 1%, 5% and 10%) and LightGBM trained on the same patient subsets (otherwise seed 42; 100%: main-text Table 2). CEHR-BERT's predictions were nearly constant (standard deviation below 0.005) at discharge 1%; readmission 1%, 5% and 10%; AKI 10% at seed 42, and in 8 of the 10 runs at seeds 142 and 242 (the other two: discharge 1%, seed 242, 0.5575; readmission 10%, seed 142, 0.5008). CEHR-BERT's seed sets the initialization of its classification head, the data order and dropout; unlike VEX-EHR's, its training/validation split stays fixed.*

| Task, fraction | VEX-EHR, seeds 42 / 142 / 242 | VEX-EHR, mean | CEHR-BERT, seeds 42 / 142 / 242 | LightGBM |
|---|---|---|---|---|
| **Discharge** | | | | |
| 1% | 0.7451 / 0.7665 / 0.7745 | 0.7620 | 0.5625 / 0.5737 / 0.5575 | 0.6480 |
| 5% | 0.8620 / 0.8548 / 0.8689 | 0.8619 | 0.6874 / 0.6111 / 0.6999 | 0.7824 |
| 10% | 0.8912 / 0.8899 / 0.8825 | 0.8879 | 0.7780 / 0.7506 / 0.7751 | 0.8053 |
| 50% | 0.9140 | 0.9140 | 0.8392 | 0.8485 |
| 100% | 0.9172 | 0.9172 | 0.8480 | 0.8680 |
| **Readmission** | | | | |
| 1% | 0.5410 / 0.5321 / 0.5668 | 0.5466 | 0.5015 / 0.4830 / 0.5112 | 0.4868 |
| 5% | 0.6275 / 0.6470 / 0.6116 | 0.6287 | 0.4734 / 0.4983 / 0.4833 | 0.5918 |
| 10% | 0.6596 / 0.6553 / 0.6623 | 0.6591 | 0.4858 / 0.5008 / 0.4600 | 0.5833 |
| 50% | 0.7024 | 0.7024 | 0.6491 | 0.6490 |
| 100% | 0.7090 / 0.7195 / 0.7174 | 0.7153 | 0.6501 | 0.6767 |
| **AKI** | | | | |
| 5% | 0.7739 / 0.7860 / 0.7749 | 0.7783 | 0.6585 / 0.5996 / 0.6645 | 0.6850 |
| 10% | 0.8049 / 0.7859 / 0.7900 | 0.7936 | 0.4648 / 0.4552 / 0.4958 | 0.7619 |
| 50% | 0.8282 | 0.8282 | 0.7489 | 0.8009 |
| 100% | 0.8424 / 0.8412 / 0.8588 | 0.8475 | 0.7914 | 0.8282 |

Fractions are of the fine-tuning pool; 1% of discharge is about 330 training instances with 43 positives and 32–41 validation instances, so epoch selection at 1% is close to random. LightGBM at 100% is the value in main-text Table 2. AUPRC at every fraction is given below (Figure S2, Table S13).

**Interpretation.** VEX-EHR's AUROC is above LightGBM's and CEHR-BERT's at every fraction on every task. On discharge, 5% of the labels (about 1,900 labeled instances) gave 0.855–0.869, above CEHR-BERT trained on all labels (0.848), and 10% gave 0.883–0.891, above LightGBM trained on all labels (0.868; main-text Table 2); on readmission, 10% of the labels gave 0.655–0.662, above CEHR-BERT with all labels (0.650), and 50% gave 0.702, above LightGBM with all labels (0.677). In paired tests, VEX-EHR was more accurate than LightGBM at every fraction of discharge (+0.066 to +0.127), at 10% and 50% of readmission (+0.053 to +0.079) and at 5% of AKI (+0.089 to +0.101), with every interval excluding zero; at 5% of readmission and at 10% and 50% of AKI, at least one interval included zero.

Figure S2 and Table S13 repeat main-text Figure 4 and Table S12 for AUPRC, the secondary metric. VEX-EHR's values are the test AUPRC of the same runs. LightGBM's were computed from its stored predictions for the same patient subsets, and CEHR-BERT's are those of its seed-42 runs on the same subsets; at 100% both are the values in main-text Table 2. AUPRC is reported descriptively; test prevalence is 12.5% (discharge), 10.9% (readmission) and 10.9% (AKI). VEX-EHR's AUPRC was above LightGBM's and CEHR-BERT's at every fraction on every task, except readmission with all labels, where seed 42 (0.259) was just below LightGBM (0.260).

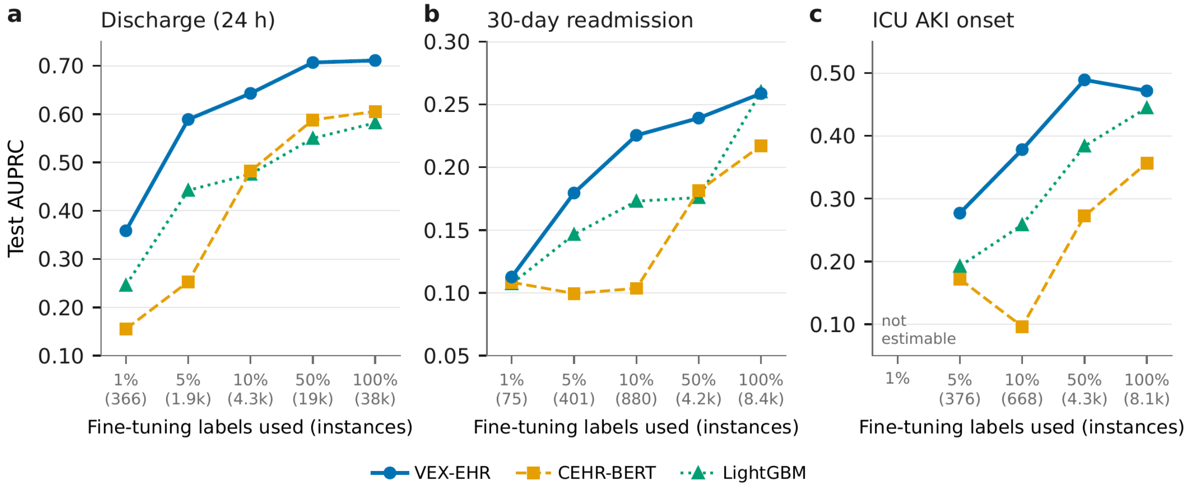

***Figure S2.*** *Test AUPRC against the fraction of fine-tuning labels used, laid out as main-text Figure 4 (a, discharge; b, readmission; c, AKI; labeled instances in parentheses; all three models fine-tuned on the same patient subsets, fine-tuning seed 42; 100%: main-text Table 2; CEHR-BERT's nearly constant runs as in Table S12). Values are in Table S13. Descriptive.*

***Table S13.*** *Label scaling: test AUPRC of VEX-EHR (seeds 42 / 142 / 242) and of CEHR-BERT and LightGBM trained on the same patient subsets (seed 42; 100%: main-text Table 2); CEHR-BERT's nearly constant runs are listed under Table S12.*

| Task, fraction | VEX-EHR, seeds 42 / 142 / 242 | CEHR-BERT | LightGBM |
|---|---|---|---|
| Discharge, 1% | 0.3585 / 0.3475 / 0.3815 | 0.1552 | 0.2457 |
| Discharge, 5% | 0.5890 / 0.5666 / 0.6022 | 0.2531 | 0.4422 |
| Discharge, 10% | 0.6430 / 0.6557 / 0.6233 | 0.4821 | 0.4757 |
| Discharge, 50% | 0.7068 | 0.5881 | 0.5499 |
| Discharge, 100% | 0.7111 | 0.6055 | 0.5819 |
| Readmission, 1% | 0.1126 / 0.1166 / 0.1195 | 0.1084 | 0.1073 |
| Readmission, 5% | 0.1795 / 0.2036 / 0.1679 | 0.0995 | 0.1465 |
| Readmission, 10% | 0.2254 / 0.2337 / 0.2295 | 0.1036 | 0.1730 |
| Readmission, 50% | 0.2392 | 0.1812 | 0.1759 |
| Readmission, 100% | 0.2587 / 0.2743 / 0.2662 | 0.2172 | 0.2601 |
| AKI, 5% | 0.2770 / 0.3053 / 0.2968 | 0.1721 | 0.1925 |
| AKI, 10% | 0.3781 / 0.3412 / 0.3585 | 0.0964 | 0.2583 |
| AKI, 50% | 0.4891 | 0.2724 | 0.3837 |
| AKI, 100% | 0.4716 / 0.4465 / 0.5016 | 0.3567 | 0.4445 |

VEX-EHR: per seed (42 / 142 / 242; one seed at 50% and for discharge at 100%). LightGBM: computed from its stored predictions (1–50%) and as in main-text Table 2 at 100%. AKI at 1% (4 patients) is not estimable.

### S11. Fine-tuning seeds

Seeds 42, 142 and 242 were fixed before any replicate ran. The seed drives head initialization, data order, dropout and the training/validation cut of the fine-tuning pool; the test rows and the continued-pretraining artifact are fixed. Seeds are spaced by 100 because each epoch reseeds with seed + epoch; consecutive seeds would share per-epoch random streams. For the main model, readmission and AKI were fine-tuned at all three seeds and discharge at seed 42 only (about 153 GPU-hours per run).

***Table S14.*** *Per-seed results of the main model on the 2019–2024 test set.*

| Task, seed | Train / val | Sel. epoch | Val AUROC | Test AUROC | Test AUPRC |
|---|---:|---:|---:|---:|---:|
| Discharge, 42 | 34,447 / 3,843 | 6 | 0.9136 | 0.9172 | 0.7111 |
| Readmission, 42 | 7,525 / 835 | 4 | 0.7112 | 0.7090 | 0.2587 |
| Readmission, 142 | 7,524 / 836 | 4 | 0.7990 | 0.7195 | 0.2743 |
| Readmission, 242 | 7,525 / 835 | 4 | 0.7083 | 0.7174 | 0.2662 |
| AKI, 42 | 7,256 / 803 | 5 | 0.8976 | 0.8424 | 0.4716 |
| AKI, 142 | 7,225 / 834 | 8 | 0.8070 | 0.8412 | 0.4465 |
| AKI, 242 | 7,262 / 797 | 4 | 0.8153 | 0.8588 | 0.5016 |

Across seeds, test AUROC varies by about 0.01 (readmission) and 0.02 (AKI), while the selected validation AUROC varies by about 0.09: the validation score reflects which patients form the small validation cut and is usable only to choose an epoch within a run. VEX-EHR-Lite's three seeds gave discharge 0.9006 / 0.8964 / 0.9001, readmission 0.7090 / 0.7119 / 0.7036 and AKI 0.8078 / 0.8251 / 0.8224 (Section S6).

### S12. Discrimination curves, calibration and threshold-dependent metrics

Figure S3 shows ROC and precision-recall curves of the main model (seed 42) on the 2019–2024 test instances, and reliability curves of its raw probabilities (deciles of predicted probability). The model was trained with class weighting (positive-class weight n_neg/n_pos), which inflates predicted probabilities: on every task it over-predicts (calibration intercepts −1.53 to −2.71). Table S15 gives the Brier score, the expected calibration error (ECE; 10 equal-width bins) and the calibration intercept and slope, before and after a logistic recalibration fitted on the validation predictions of each fine-tuning run and applied unchanged to the test instances. Recalibration brings the ECE to 0.010–0.033 and the slopes to 0.80–1.01, without changing discrimination; the AKI slope is furthest from 1 because the AKI validation set is small (803 instances).

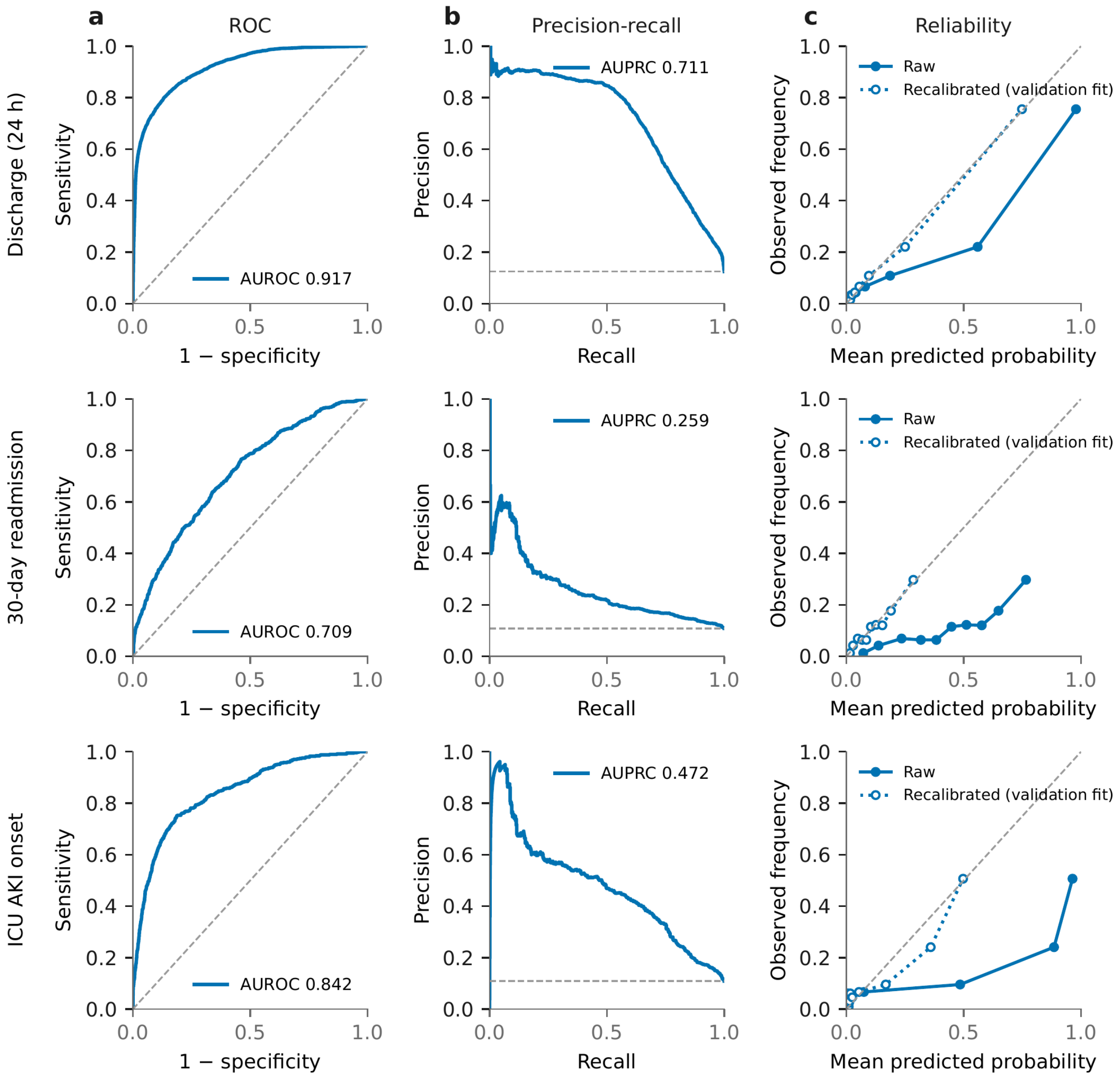

***Figure S3.*** *ROC (a), precision-recall (b) and reliability curves (c; deciles of predicted probability) for VEX-EHR (main model, seed 42) on the 2019–2024 test instances. Reliability: filled markers, raw probabilities; open markers, after the logistic recalibration fitted on the validation set (Table S15). Dashed lines: chance (ROC), prevalence (precision-recall), perfect calibration (reliability).*

***Table S15.*** *Calibration of VEX-EHR before (raw) and after logistic recalibration (fitted on each fine-tuning run's validation predictions, applied to the test instances). Brier score / ECE; calibration intercept / slope.*

| Task, model | Raw: Brier / ECE | Raw: intercept / slope | Recalibrated: Brier / ECE / slope |
|---|---|---|---|
| Discharge, VEX-EHR | 0.079 / 0.065 | −1.53 / 0.61 | 0.059 / 0.012 / 1.01 |
| Readmission, VEX-EHR | 0.202 / 0.301 | −2.03 / 0.82 | 0.090 / 0.010 / 0.96 |
| AKI, VEX-EHR | 0.155 / 0.166 | −2.71 / 0.39 | 0.077 / 0.033 / 0.80 |

ECE, expected calibration error (10 equal-width bins, weighted by bin size). Across the three fine-tuning seeds, VEX-EHR's raw ECE ranged 0.17–0.30 (readmission) and 0.11–0.33 (AKI); discharge has one seed.

***Table S16.*** *Threshold-dependent metrics of VEX-EHR (main model, seed 42), at a fixed threshold of 0.5 for precision, recall, F1 and accuracy. The model is not calibrated (it is trained with class-weighted loss), so these four metrics describe one operating point, not discrimination.*

| Task | Model | AUROC | AUPRC | Precision | Recall | F1 | Accuracy |
|---|---|---|---|---|---|---|---|
| Discharge | VEX-EHR (seed 42) | 0.9172 | 0.7111 | 0.5804 | 0.7224 | 0.6437 | 0.8999 |
| Readmission | VEX-EHR (seed 42) | 0.7090 | 0.2587 | 0.1861 | 0.6313 | 0.2875 | 0.6602 |
| AKI | VEX-EHR (seed 42) | 0.8424 | 0.4716 | 0.3300 | 0.7496 | 0.4582 | 0.8064 |

### S13. Subgroups and fairness

Discrimination was examined within subgroups defined by sex and by age at the prediction time (<45, 45–64 and ≥65 years; bands commonly used in US health statistics, which also gave groups of similar size; every test patient was 18 or older), the demographic fields available in the extract; race is analyzed below. Age is the patient's recorded age at the latest visit on or before the prediction time. Sex and age were known for at least 99.8% of test rows on every task. Table S17 gives the AUROC of VEX-EHR, LightGBM and CEHR-BERT in each subgroup, on the same instances. These analyses are exploratory and not adjusted for multiple comparisons. Discharge AUROC was at least 0.895 in every subgroup. Readmission AUROC was higher in women (0.738) than in men (0.669) and decreased with age (0.763, 0.690 and 0.650). AKI AUROC was lowest in patients aged 65 or older (0.793). LightGBM and CEHR-BERT showed gaps in the same direction, and VEX-EHR's gap (female minus male; younger than 45 minus 65 or older) was not significantly larger than either comparator's on any task (difference of gaps, patient-clustered bootstrap, 2,000 draws); on discharge its age gap was smaller (0.033, against

0.053 for LightGBM and 0.060 for CEHR-BERT; differences −0.020, 95% CI −0.036 to −0.004, and −0.027, −0.045 to −0.010). VEX-EHR had the highest AUROC of the three models in every group.

***Table S17.*** *Discrimination by sex and age on the 2019–2024 test instances: AUROC of VEX-EHR (main model, seed 42; for readmission and AKI also the mean over seeds 42, 142 and 242), LightGBM and CEHR-BERT.*

| Task, subgroup | Rows / patients / positives | AUROC, VEX-EHR (seed 42) | VEX-EHR, mean of seeds | LightGBM | CEHR-BERT |
|---|---|---|---|---|---|
| Discharge, all | 26,300 / 3,345 / 3,292 | 0.917 | — | 0.868 | 0.848 |
| Discharge, female | 13,337 / 1,905 / 1,991 | 0.929 | — | 0.883 | 0.865 |
| Discharge, male | 12,955 / 1,439 / 1,301 | 0.897 | — | 0.842 | 0.819 |
| Discharge, age <45 | 8,550 / 1,358 / 1,386 | 0.928 | — | 0.887 | 0.876 |
| Discharge, age 45–64 | 9,081 / 983 / 924 | 0.920 | — | 0.870 | 0.834 |
| Discharge, age ≥65 | 8,669 / 1,043 / 982 | 0.895 | — | 0.834 | 0.816 |
| Readmission, all | 5,471 / 3,537 / 594 | 0.709 | 0.715 | 0.677 | 0.650 |
| Readmission, female | 2,977 / 1,988 / 259 | 0.738 | 0.740 | 0.691 | 0.667 |
| Readmission, male | 2,493 / 1,548 / 335 | 0.669 | 0.680 | 0.651 | 0.628 |
| Readmission, age <45 | 2,028 / 1,424 / 184 | 0.763 | 0.765 | 0.724 | 0.703 |
| Readmission, age 45–64 | 1,697 / 1,042 / 240 | 0.690 | 0.698 | 0.657 | 0.631 |
| Readmission, age ≥65 | 1,746 / 1,111 / 170 | 0.650 | 0.663 | 0.629 | 0.583 |
| AKI, all | 5,226 / 392 / 571 | 0.842 | 0.847 | 0.828 | 0.791 |
| AKI, female | 2,295 / 162 / 228 | 0.855 | 0.863 | 0.854 | 0.784 |
| AKI, male | 2,919 / 229 / 343 | 0.832 | 0.835 | 0.810 | 0.798 |
| AKI, age <45 | 1,289 / 90 / 144 | 0.886 | 0.879 | 0.854 | 0.854 |
| AKI, age 45–64 | 2,279 / 158 / 236 | 0.849 | 0.864 | 0.846 | 0.817 |
| AKI, age ≥65 | 1,658 / 145 / 191 | 0.793 | 0.789 | 0.784 | 0.695 |

A patient whose age crosses a boundary contributes rows to two age groups. Discharge uses the 26,300 rows VEX-EHR scored.

**Race.** Race is the recorded race in the institution's OMOP person table, linked by patient identifier (matched for 99.97% of 2019–2024 test patients and 99.93% of 2025 patients; 77.1% and 76.2% White). As in an earlier comparison at this institution (main-text reference 7), patients are grouped as White and non-White, with unknown and declined race counted as non-White; a sensitivity analysis excludes them. Table S18 gives VEX-EHR's AUROC in the two groups and its race gap (non-White minus White) on the 2019–2024 test set and the 2025 cohort (all, new and returning patients), with patient-clustered bootstrap 95% intervals over the whole cohort (2,000 draws), per seed (readmission and AKI: seeds 42, 142 and 242; discharge: seed 42). VEX-EHR's AUROC was not significantly lower in non-White than in White patients in any cohort or at any seed: no negative gap excluded zero. It was significantly higher in non-White patients on discharge in 2025 (all and new patients) and on readmission in 2025 (all, new and returning patients, at every seed). Excluding unknown and declined race gave the same conclusions.

***Table S18.*** *AUROC of VEX-EHR by race: AUROC in White / non-White patients and the race gap (non-White − White), per seed.*

| Task, cohort | AUROC, White / non-White | Race gap, seeds 42 / 142 / 242 |
|---|---|---|
| Discharge, 2019–2024 | 0.915 / 0.925 | +0.011 |
| Discharge, 2025, all | 0.960 / 0.971 | +0.011* |
| Discharge, 2025, new | 0.963 / 0.975 | +0.012* |
| Discharge, 2025, returning | 0.954 / 0.959 | +0.006 |
| Readmission, 2019–2024 | 0.713 / 0.722 | +0.008 / +0.016 / +0.003 |
| Readmission, 2025, all | 0.691 / 0.740 | +0.048* / +0.050* / +0.049* |
| Readmission, 2025, new | 0.702 / 0.736 | +0.034* / +0.036* / +0.032* |
| Readmission, 2025, returning | 0.641 / 0.686 | +0.045* / +0.044* / +0.046* |
| AKI, 2019–2024 | 0.846 / 0.847 | −0.006 / −0.005 / +0.017 |
| AKI, 2025, all | 0.795 / 0.825 | +0.031 / +0.029 / +0.028 |
| AKI, 2025, new | 0.787 / 0.817 | +0.032 / +0.025 / +0.031 |
| AKI, 2025, returning | 0.812 / 0.833 | +0.022 / +0.026 / +0.016 |

AUROC: White / non-White, mean of seeds. Gaps: seeds 42 / 142 / 242 (discharge: 42). * Patient-clustered 95% interval excludes zero. Non-White includes unknown and declined race.

**Thresholds.** The validation Youden threshold (maximum TPR − FPR) moved widely between VEX-EHR's fine-tuning seeds, whose validation sets also differ: readmission 0.645, 0.477 and 0.307; AKI 0.855, $3.8\times10^{-5}$ and 0.804, the seed-142 AKI model's scores lying close to zero (median validation score $4\times10^{-5}$). In 2025 the three AKI models flagged 17.6%, 43.6% and 38.9% of instances. Discharge: 0.234. A decision threshold would therefore need to be set locally, on the deployed model, before use.

### S14. Vocabulary map

Each vocabulary embedding of the backbone was projected to two dimensions: 20,000 frequent English text tokens (the most frequent WikiText-2 training tokens that contain a letter) and the 17,601 added codes. One UMAP projection was fitted jointly on the text-token embeddings, the semantic initialization and every configuration's trained code embeddings, so positions compare across panels (main-text Figure 5). Table S19 gives the corresponding measurements in the full 3,840-dimensional space. Fine-tuning does not change the embedding table (it trains LoRA and the head only), so these are the tables the fine-tuned models use. The configurations at $2\times10^{-5}$ (the 1,024-token reference, VEX-EHR-Lite and CPT sliding 2,048 at $2\times10^{-5}$ + FT 4,096) changed their code embeddings by less than 0.01 in cosine. The main model, at $3\times10^{-4}$, moved them substantially (mean cosine 0.564 to their initialization) and away from the text tokens, while the embeddings of its original vocabulary stayed nearly fixed (0.998 over all 262,144 text tokens; 0.996 over the 20,000 in Table S19). Same-type structure was already strong after semantic initialization (0.974) and absent after random initialization (0.286); continued pretraining at $2\times10^{-5}$ or from random initialization left it nearly unchanged, and the main model's raised it to 0.991. Main-text Figure 5c and d give each token's angle between its embedding before and after continued pretraining (median, interquartile range): 56.6° (52.8–59.3°) for the 17,601 codes and 2.7° (1.9–3.8°) for the 262,144 text tokens in the main model, and 4.3° (3.6–5.2°) for the randomly initialized codes.

***Table S19.*** *Code and text-token embeddings after continued pretraining, by configuration (all 17,601 codes; 20,000 text tokens).*

| Configuration | Cosine to initialization: codes / text | Cosine with the nearest text token: before → after | Same-type share of the 10 nearest codes: before → after |
|---|---|---|---|
| 1,024-token reference | 0.998 / 0.999 | 0.453 → 0.453 | 0.974 → 0.974 |
| Random initialization (1,024) | 0.997 / 1.000 | 0.064 → 0.065 | 0.286 → 0.301 |
| VEX-EHR-Lite (2,048) | 0.998 / 0.999 | 0.453 → 0.453 | 0.974 → 0.974 |
| CPT sliding 2,048 at $2\times10^{-5}$ + FT 4,096 (two epochs) | 0.995 / 0.999 | 0.453 → 0.450 | 0.974 → 0.975 |
| Main model (full-history corpus, $3\times10^{-4}$, one epoch) | 0.564 / 0.996 | 0.453 → 0.230 | 0.974 → 0.991 |

Means over tokens. Cosine to initialization: each embedding with its value before continued pretraining. Nearest text token: highest cosine between a code embedding and any of the 20,000 text-token embeddings. Same-type share: among each code's 10 nearest codes (cosine), the fraction of its own type (diagnosis, medication, procedure, laboratory, audit-log).

### S15. Training-free probe: can a code token be read back into English?

**Design.** 500 codes (100 each of diagnoses, medications, procedures, laboratory orders and audit-log actions; within each, 50 above and 50 at or below the category's median frequency in the pretraining corpus), plus a harder set of 200 (diagnoses sharing the same three-character ICD-10 category; medications sharing the ingredient). Each item offers 10 candidate descriptions: the code's own (the text its embedding was initialized from) and 9 distractors of the same category, distinct in content (for medications, descriptions differing only in an application number or brand name count as the same). The prompt is “Medical code: `<code>` Meaning: `<candidate>`”. A candidate is scored by the mean log-probability of its tokens, with the softmax restricted to the backbone’s 262,144 text tokens. The model chooses the highest-scoring candidate (chance 10%). No model is trained. Conditions: the backbone with semantic initialization and no continued pretraining (M0); VEX-EHR-Lite (Section S6) after continued pretraining with its LoRA on (M1) and off, i.e., its trained embedding table read by the original transformer (M3); the 1,024-token model (M1′); and the randomly initialized 1,024-token model after the same continued pretraining (M2). Item-level bootstrap, 2,000 draws, seed 42.

**Scoring.** Under the mean log-probability, every condition was near chance (M0 10.2%, M1 15.6%, M2 7.4%), because each candidate’s prior likelihood dominated the score. We therefore report the pointwise mutual information (PMI), score(candidate | code) − score(candidate | no code), which removes that prior.

***Table S20.*** *Probe accuracy (top-1 among 10, %) under the PMI score.*

| Condition | All | ICD-10 | Drug | Proc. | Lab | Audit | Frequent | Rare | Hard set |
|---|---|---|---|---|---|---|---|---|---|
| M0 semantic init, no continued pretraining | 26.6 | 24 | 20 | 17 | 13 | 59 | 26.4 | 26.8 | 15.0 |
| M1 VEX-EHR-Lite, LoRA on | 39.8 | 49 | 25 | 21 | 29 | 75 | 45.2 | 34.4 | 20.5 |
| M3 VEX-EHR-Lite, LoRA off (table only) | 27.8 | 26 | 21 | 21 | 15 | 56 | 28.0 | 27.6 | 18.5 |
| M1′ 1,024-token model, LoRA on | 36.2 | 40 | 20 | 28 | 23 | 70 | 41.6 | 30.8 | 17.5 |
| M2 random init, same pretraining | 8.6 | 6 | 5 | 15 | 5 | 12 | 8.4 | 8.8 | 12.0 |

Paired contrasts (main set, percentage points, 95% interval; without audit-log items in brackets): M0 − chance +16.6 (+12.8 to +20.6) [+8.5]; M3 − M0 +1.2 (−1.4 to +3.8) [+2.2, interval includes 0]; M1 − M0 +13.2 (+8.6 to +17.8) [+12.5]; M2 − chance −1.4 (−3.8 to +1.0); M1′ − M2 +27.6 (+23.0 to +32.0) [+20.0]; M1 − M1′ +3.6 (−0.4 to +8.0). The LoRA gain (M1 − M3) is +17.2 for frequent and +6.8 for rare codes. Audit-log descriptions are short English phrases and are the easiest. One model per condition and one prompt template. Readability does not show that the downstream benefit of semantic initialization comes from this meaning.

**Interpretation.** The semantic initialization gives each code an English anchor that the backbone can partly read. Continued pretraining leaves it in the embedding table unchanged, and the adapter learned during continued pretraining reads it better, most for codes seen often. Continued pretraining alone, from a random initialization, does not produce it.

### S16. AKI: predictions whose input already contains an AKI diagnosis

Table S21 gives the AKI sensitivity analysis described in the main text (Methods and Discussion): the 2019–2024 AKI test predictions, split by whether the input already contained an AKI diagnosis code.

***Table S21.*** *AKI test predictions (2019–2024) by whether the 24-hour input already contained an AKI diagnosis code (ICD-10 N17.0–N17.9). The trained models were re-evaluated on each subset without retraining: VEX-EHR at fine-tuning seeds 42, 142 and 242, and LightGBM. An AKI diagnosis code was in the input of 211 of 571 positive (37.0%) and 373 of 4,655 negative (8.0%) predictions; used alone as a predictor, its AUROC was 0.645. Difference: VEX-EHR minus LightGBM AUROC, with a paired patient-clustered bootstrap 95% CI (2,000 resamples).*

| Test predictions | Predictions (positive); patients | VEX-EHR AUROC, seeds 42 / 142 / 242 | LightGBM AUROC | Difference (95% CI), seeds 42 / 142 / 242 |
|---|---|---|---|---|
| All | 5,226 (571); 392 | 0.842 / 0.841 / 0.859 | 0.828 | +0.014 (−0.019 to 0.045); +0.013 (−0.020 to 0.044); +0.031 (−0.003 to 0.064) |
| No AKI diagnosis code in input | 4,642 (360); 367 | 0.830 / 0.830 / 0.848 | 0.804 | +0.027 (−0.015 to 0.069); +0.026 (−0.015 to 0.068); +0.045 (0.004 to 0.085) |
| AKI diagnosis code in input | 584 (211); 92 | 0.735 / 0.737 / 0.756 | 0.744 | −0.009 (−0.087 to 0.076); −0.007 (−0.083 to 0.078); +0.011 (−0.074 to 0.089) |
